# Intradermal versus Intramuscular Administration of Influenza Vaccination: Rapid Review and Meta-analysis

**DOI:** 10.1101/2020.10.06.20205989

**Authors:** Oluwaseun Egunsola, Fiona Clement, John Taplin, Liza Mastikhina, Joyce W. Li, Diane L. Lorenzetti, Laura E. Dowsett, Tom Noseworthy

## Abstract

**Background:** Vaccinations are essential for prevention of influenza. We synthesized the published literature on the immunogenicity and safety of the influenza vaccine at reduced or full intradermal doses compared with full intramuscular doses.

**Methods:** A rapid review of the literature was completed. MEDLINE, EMBASE, and the Cochrane Central Register of Controlled Trials were searched for studies published from 2010 until June 5^th^, 2020. All studies comparing intradermal and intramuscular influenza vaccinations were included. Random-effects meta-analyses of immunogenicity and safety outcomes were conducted.

**Results:** A total of 30 relevant studies were included. Seroconversion rates were equivalent between the 3 mcg, 6 mcg, 7.5 mcg, and 9 mcg intradermal vaccine doses and the 15 mcg intramuscular vaccine dose for each of the H1N1, H3N2, and B strains, but significantly higher with the 15 mcg intradermal compared with the 15 mcg intramuscular dose, for the H1N1 (RR 1.10, 95% CI: 1.01-1.20) and B strains (RR 1.40, 95% CI: 1.13-1.73). Seroprotection rates for the 9 mcg and 15 mcg intradermal doses were equivalent with the 15 mcg intramuscular dose for all the three strains, except for the 15 mcg intradermal dose for the H1N1 strain which was significantly higher (RR 1.05, 95% CI: 1.01-1.09). Local adverse events were significantly higher with intradermal doses. Fever and chills were significantly higher with the 9 mcg intradermal dose, while all other systemic adverse events were equivalent for all doses.

**Conclusion:** Reduced dose intradermal influenza vaccination appears to be a reasonable alternative to standard dose intramuscular vaccination because of the similarity in immunogenicity.

## 1 Background

Influenza infection causes three-to-five million severe illnesses and approximately half a million annual deaths globally.^1^ It is a highly contagious disease characterized by high fever, cough, sore throat, headache, chills, lack of appetite, and fatigue.^2^ Vaccinations are essential for prevention of influenza and can be administered intradermally or intramuscularly, with the latter being the more common method.^3^

The interest in intradermal vaccines has been increasing as a solution to mitigate potential vaccine shortages, which could occur from unanticipated loss of expected supplies or from excessive demand due to high rates of infection, such as during pandemics.^4^ With the approval of new intradermal vaccines,^2 5 6^ new delivery devices, including mini-needles, microneedles, patches and disposable-syringe jet injectors, have become available.^3 7^

Intradermal vaccinations have a dose-sparing effect;^3^ therefore, smaller doses of intradermal vaccines may be sufficient to produce an antigenic response that is similar to standard intramuscular doses. The dermis is rich in dendritic cells, which are very potent antigen-presenting cells that elicit cell mediated immune responses, especially CD4+ and CD8+ T-cell responses which are essential in the immune response to influenza viruses.^4 8^ Intramuscular injection bypasses this immune system response and delivers the vaccine directly into the muscular tissue, which has relatively few resident antigen-presenting cells.^9^ Previous studies have compared the immunogenicity (ability of the vaccine to evoke an immune response) and safety of intradermal and intramuscular influenza vaccines; however, the magnitude of the effect has not yet been examined. In this study, we synthesized the published literature on the immunogenicity and safety of the influenza vaccine at reduced or regular intradermal doses compared with a full intramuscular dose.

## 2 Methods

### 2.1 Literature Search

A rapid review of the literature was completed. MEDLINE, EMBASE, and the Cochrane Central Register of Controlled Trials were searched for studies published from inception until June 5^th^, 2020. Terms aimed to capture the technology of interest, such as “intradermal,” “ID injection” and “mantoux” were combined using the Boolean Operator “and” with influenza terms. These terms were searched as text words in titles and key word headings and as MeSH subject headings when applicable. The search excluded case reports, editorials, letters, and animal studies. The search strategy was developed by a research librarian, and PRESS-reviewed by another research librarian.^10^ This search was supplemented by scrutinizing the reference lists of systematic literature reviews to ensure that all studies meeting the inclusion criteria were captured.

### 2.2 Literature Selection

Abstracts identified through database searching were screened by a single reviewer; all abstracts included at this stage proceeded to full-text review. Full-text publications were screened by a single reviewer. Publications were included if they met all inclusion criteria and failed to meet any exclusion criteria outlined in Table 1.

**Table 1.** Inclusion and Exclusion Criteria

| Inclusion Criteria | Exclusion Criteria |
| --- | --- |
| All ages, humans only<br>Must compare intradermal administration of the inactivated influenza vaccine (trivalent standard-dose influenza vaccine, quadrivalent standard-dose influenza vaccine, adjuvanted trivalent influenza vaccine, high-dose trivalent influenza vaccine) with intramuscular administration of the influenza vaccine<br>Includes one of the following:<br>lab-confirmed influenza infection<br>influenza-like illness<br>hospitalization<br>ICU admission<br>pneumonia<br>mortality<br>immunogenicity<br>adverse events<br>Randomized controlled trials (RCTs), NRCTs (e.g., such as quasi-RCTs, non-randomized trials, interrupted time series, controlled before after), and observational studies (e.g., cohort, case control)<br>Published 2010-2020 | Animal models<br>Non-influenza vaccination<br>Whole virus vaccinations<br>Inclusion of immune compromised patients<br>Study designs without a control or comparator such as: cross-sectional, case series, case reports, and qualitative studies.<br>Commentaries, conference proceedings |

### 2.3 Data Extraction

For all included studies, year of publication, country, study design, dates of recruitment, study inclusion and exclusion criteria, setting, patient characteristics, treatment protocol (e.g., intention-to-treat, per-protocol), sample size, follow-up time, geometric mean titer (GMT), defined as the antilog of the arithmetic mean of the log-transformed titers, seroconversion and seroprotection rates and all relevant outcomes were extracted by a single reviewer and verified by a second reviewer using standardized data extraction forms. Discrepancies between reviewers during data extraction were resolved through consensus.

### 2.4 Quality Assessment

The quality of randomized controlled trials were assessed using the Cochrane Handbook Risk of Bias Assessment Tool (version 5.1.0).^11^ Each study was assessed using seven criteria broadly covering the areas of randomization, allocation concealment, blinding of participants and personnel, blinding of outcome assessment, incomplete outcome data, and selective reporting. Each criterion was assigned a rating of “low,” “high,” or “unclear.”

The quality of cohort studies was assessed using the Newcastle Ottawa Scale. Each study was assessed across three categories: selection, comparability, and outcome. Items within selection and comparability were assigned up to one ‘star’ for high quality, while items within comparability were assigned a maximum of two ‘stars.’

Quality assessment was completed by a single reviewer and verified by a second reviewer. Discrepancies were resolved through discussion. Studies were not excluded based on quality assessment.

### 2.5 Data Analysis

Random-effects meta-analysis was conducted, utilizing the DerSimonian and Laird estimator^12^ for Tau. Statistical heterogeneity was assessed using the I^2^ measure, with values above or below 50% considered high and low heterogeneity respectively. A continuity correction of 0.5 was used where appropriate, allowing the inclusion of zero-total event trials.^13^ Stratified analyses by dose were completed for the geometric mean titer, seroconversion, seroprotection, and adverse events. Only immunogenicity outcomes for days 21-30 post-vaccination were analyzed. Subgroup analyses of immunogenicity outcomes were conducted for studies involving participants ≥60 years of age. Risk ratios were calculated for categorical outcomes and the ratio of geometric means calculated for GMT, as described by Friedrich el al.^14^ Publication bias for small studies with missing small effect sizes was assessed using Egger’s test^15^ when the number of studies was greater than four, and, where appropriate, the Duval & Tweedie’s trim-and-fill method^16^ was used to adjust for funnel plot asymmetry. All analyses were completed in R version 3.6.1.

## 3 Results

### 3.1 Study Characteristics

The search strategy yielded 914 unique citations; 869 were excluded after deduplication and abstract review. Forty-five studies proceeded to full-text review (Figure 1). Fifteen studies were excluded for the following reasons: study design (n=5); incorrect outcome (n=5); duplicate (n=2); incorrect study population (n=1); and publication year not of interest (n=1). A total of 30 relevant studies were included in the final dataset (Figure 1).

**Figure 1.**
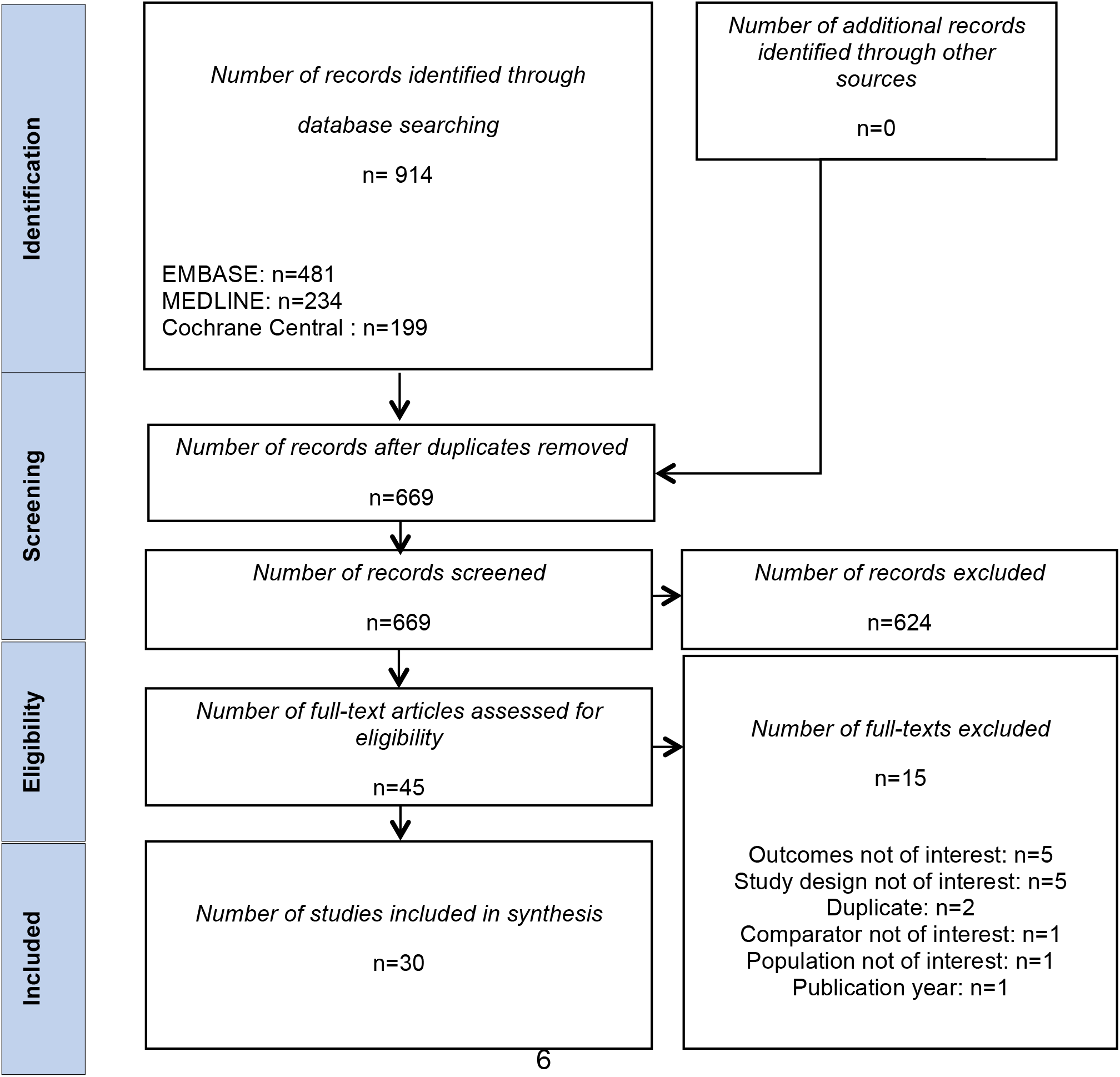
PRISMA Flowchart of Included and Excluded Studies

Twenty-nine of the studies were randomized controlled trials involving a total of 13,759 participants,^17-45^ and one study was a cohort study of 164,021 participants.^46^ All studies were published between 2010 and 2019. Sixteen studies were multi-centre,^17-19 21 22 28 30 32 33 36 37 40 42 43 46^ 12 were single-centre,^23-27 29 31 34 35 38 39 44^ and two studies did not report the setting.^20 41^ Half of the studies involved only participants ≥60 years old or reported data for participants ≥60 years old.^17 18 20 21 23-26 28 33 40 42 43 45 46^

### 3.2 Quality Assessment

The majority of the studies had some bias stemming from the randomization process; six studies were at low risk of bias;^19 22 23 29 31 34^ and two studies were at high risk.^27 33^ All but two low risk studies^22 34^ had some risk of bias due to deviations from intended interventions. All the included studies had low risk of bias due to missing outcome data. All but one high risk study^33^ were of low risk of bias stemming from the measurement of outcomes. Lastly, all the studies were of some concern of bias regarding selection of the reported results. Overall, all the studies except two high risk studies,^27 33^ were of some concern for bias.

The only included cohort study was allocated nine stars.^46^ It was judged to be representative of the exposed population. Exposure and outcomes were ascertained from secure records and record linkage respectively. The cohorts were comparable and follow-up was long and adequate.

### 3.3 Meta-analysis

#### 3.3.1 Seroconversion

Seroconversion rates were equivalent between the 3 mcg, 6 mcg, 7.5 mcg, and 9 mcg intradermal vaccine doses and the 15mcg intramuscular vaccine dose for each of the H1N1, H3N2, and B strains. Furthermore, seroconversion rate for the H3N2 strain was also equivalent between the 15 mcg intradermal and 15 mcg intramuscular doses, but significantly higher with the 15 mcg intradermal compared with 15 mcg intramuscular dose, for the H1N1 (RR 1.10, 95% CI: 1.01-1.20) and B strains (RR 1.40, 95% CI: 1.13-1.73) (Table 2).

**Table 2.**
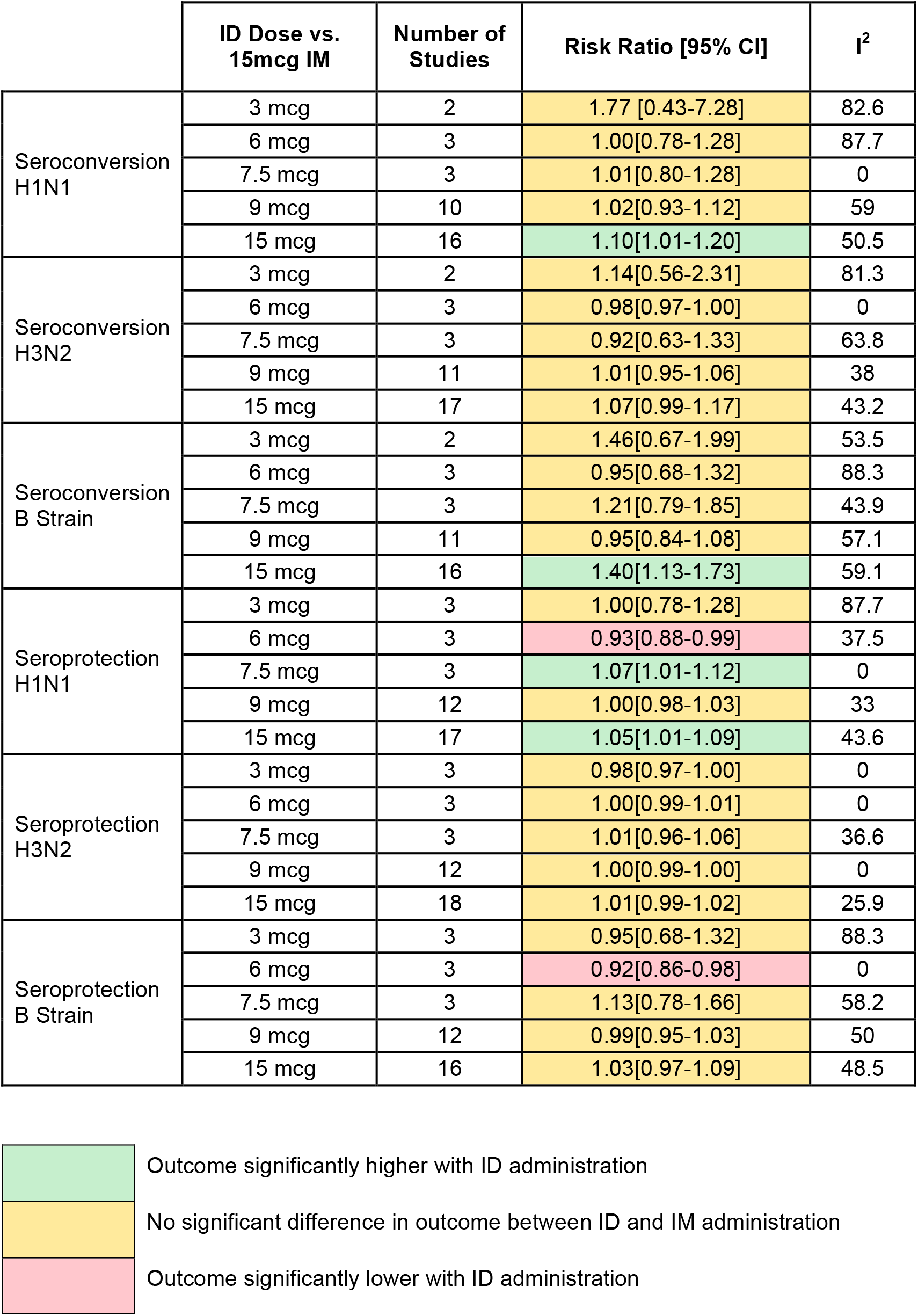
Seroconversion and Seroprotection of ID Doses versus Standard IM Dose

|  | ID Dose vs.<br>15mcg IM | Number of<br>Studies | Risk Ratio [95% CI] | I<sup>2</sup> |
| --- | --- | --- | --- | --- |
| Seroconversion<br>H1N1 | 3 mcg | 2 | 1.77 [0.43-7.28] | 82.6 |
|  | 6 mcg | 3 | 1.00[0.78-1.28] | 87.7 |
|  | 7.5 mcg | 3 | 1.01[0.80-1.28] | 0 |
|  | 9 mcg | 10 | 1.02[0.93-1.12] | 59 |
|  | 15 mcg | 16 | 1.10[1.01-1.20] | 50.5 |
| Seroconversion<br>H3N2 | 3 mcg | 2 | 1.14[0.56-2.31] | 81.3 |
|  | 6 mcg | 3 | 0.98[0.97-1.00] | 0 |
|  | 7.5 mcg | 3 | 0.92[0.63-1.33] | 63.8 |
|  | 9 mcg | 11 | 1.01[0.95-1.06] | 38 |
|  | 15 mcg | 17 | 1.07[0.99-1.17] | 43.2 |
| Seroconversion<br>B Strain | 3 mcg | 2 | 1.46[0.67-1.99] | 53.5 |
|  | 6 mcg | 3 | 0.95[0.68-1.32] | 88.3 |
|  | 7.5 mcg | 3 | 1.21[0.79-1.85] | 43.9 |
|  | 9 mcg | 11 | 0.95[0.84-1.08] | 57.1 |
|  | 15 mcg | 16 | 1.40[1.13-1.73] | 59.1 |
| Seroprotection<br>H1N1 | 3 mcg | 3 | 1.00[0.78-1.28] | 87.7 |
|  | 6 mcg | 3 | 0.93[0.88-0.99] | 37.5 |
|  | 7.5 mcg | 3 | 1.07[1.01-1.12] | 0 |
|  | 9 mcg | 12 | 1.00[0.98-1.03] | 33 |
|  | 15 mcg | 17 | 1.05[1.01-1.09] | 43.6 |
| Seroprotection<br>H3N2 | 3 mcg | 3 | 0.98[0.97-1.00] | 0 |
|  | 6 mcg | 3 | 1.00[0.99-1.01] | 0 |
|  | 7.5 mcg | 3 | 1.01[0.96-1.06] | 36.6 |
|  | 9 mcg | 12 | 1.00[0.99-1.00] | 0 |
|  | 15 mcg | 18 | 1.01[0.99-1.02] | 25.9 |
| Seroprotection<br>B Strain | 3 mcg | 3 | 0.95[0.68-1.32] | 88.3 |
|  | 6 mcg | 3 | 0.92[0.86-0.98] | 0 |
|  | 7.5 mcg | 3 | 1.13[0.78-1.66] | 58.2 |
|  | 9 mcg | 12 | 0.99[0.95-1.03] | 50 |
|  | 15 mcg | 16 | 1.03[0.97-1.09] | 48.5 |

|  |  |
| --- | --- |
|  | Outcome significantly higher with ID administration |
|  | No significant difference in outcome between ID and IM administration |
|  | Outcome significantly lower with ID administration |

#### 3.3.2 Seroprotection

Seroprotection rates were significantly lower with the 6 mcg intradermal dose for the H1N1 (RR 0.93, 95% CI: 0.88-0.99) and the B strains (RR 0.92, 95% CI: 0.86-0.98). For the 9 mcg intradermal doses, seroprotection rates were equivalent with the 15 mcg intramuscular dose for all the three strains. The 15 mcg intradermal and 15 mcg intramuscular doses were also equivalent for H3N2 and B strains, however, seroprotection rate was significantly higher for the H1N1 strain (RR 1.05, 95% CI:1.01-1.09) (Table 2).

#### 3.3.3 Geometric Mean Titer

The GMTs were equivalent between the 3 mcg and 6 mcg intradermal doses and the 15 mcg intramuscular dose for the three strains, except for a significant decrease for H1N1 observed with the 6 mcg intradermal dose (RR 0.88, 95% CI: 0.85-0.90). Similarly, GMTs were equivalent for the H1N1 and B strains when the 9 mcg intradermal dose was compared with the 15 mcg intramuscular dose, but significantly higher for the H3N2 strain (RR 1.08, 95% CI: 1.05-1.12). The 15 mcg intradermal dose showed equivalence with the 15 mcg intramuscular dose for the H1N1 and the H3N2 strains. However, the 15 mcg intradermal dose was associated with significantly higher GMT for the B strain (RR 1.21, 95% CI: 1.11-1.32) (Table 3).

**Table 3.**
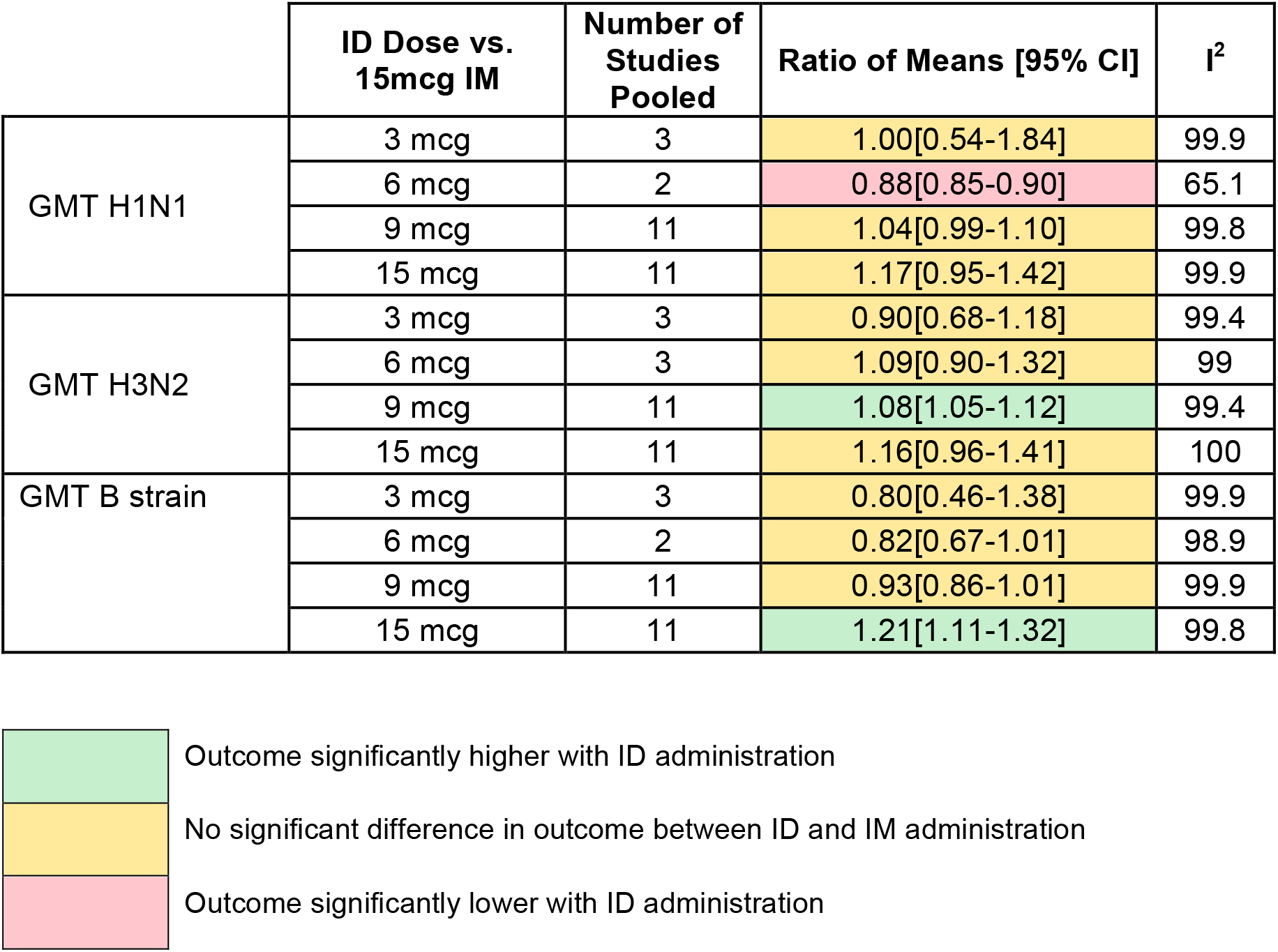
Geometric Mean Titer of ID Doses versus Standard IM Dose

|  | ID Dose vs.<br>15mcg IM | Number of<br>Studies<br>Pooled | Ratio of Means [95% CI] | I<sup>2</sup> |
| --- | --- | --- | --- | --- |
| GMT H1N1 | 3 mcg | 3 | 1.00[0.54-1.84] | 99.9 |
|  | 6 mcg | 2 | 0.88[0.85-0.90] | 65.1 |
|  | 9 mcg | 11 | 1.04[0.99-1.10] | 99.8 |
|  | 15 mcg | 11 | 1.17[0.95-1.42] | 99.9 |
| GMT H3N2 | 3 mcg | 3 | 0.90[0.68-1.18] | 99.4 |
|  | 6 mcg | 3 | 1.09[0.90-1.32] | 99 |
|  | 9 mcg | 11 | 1.08[1.05-1.12] | 99.4 |
|  | 15 mcg | 11 | 1.16[0.96-1.41] | 100 |
| GMT B strain | 3 mcg | 3 | 0.80[0.46-1.38] | 99.9 |
|  | 6 mcg | 2 | 0.82[0.67-1.01] | 98.9 |
|  | 9 mcg | 11 | 0.93[0.86-1.01] | 99.9 |
|  | 15 mcg | 11 | 1.21[1.11-1.32] | 99.8 |

#### 3.3.4 Immunogenicity in the Elderly

Subgroup analyses for immunogenicity in the elderly (aged 60 years and older) showed equivalence between the 9 mcg intradermal and 15 mcg intramuscular doses, with respect to seroconversion, seroprotection, and GMT for each of the three strains. Seroprotection rates were also equivalent between the 15 mcg intradermal and intramuscular doses for the three strains, while seroconversion rate and GMT were significantly higher with the 15 mcg intradermal dose compared with the 15 mcg intramuscular dose for the B strain (RR 1.41, 95% CI: 1.13-1.75 and RR 1.19, 95% CI: 1.09-1.30, respectively) (Table 4).

**Table 4.** Immunogenicity of ID Doses versus Standard IM Dose among Participants ≥60 Years

|  | ID Dose vs.<br>15mcg IM | Number of<br>Studies<br>Pooled | Risk Ratio/*RoM [95%<br>CI] | I<sup>2</sup> |
| --- | --- | --- | --- | --- |
| Seroconversion<br>H1N1 | 9 mcg | 2 | 1.01[0.58-1.77] | 87 |
|  | 15 mcg | 13 | 1.11[1.00-1.24] | 57.3 |
| Seroconversion<br>H3N2 | 9 mcg | 2 | 1.02[0.83-1.25] | 0 |
|  | 15 mcg | 14 | 1.12[1.00-1.25] | 52.1 |
| Seroconversion B<br>strain | 9 mcg | 2 | 1.00 [0.60-1.67] | 0 |
|  | 15 mcg | 13 | 1.41[1.13-1.75] | 59 |
| Seroprotection H1N1 | 9 mcg | 4 | 0.98[0.88-1.09] | 24.1 |
|  | 15 mcg | 14 | 1.04[1.00-1.09] | 55.1 |
| Seroprotection H3N2 | 9 mcg | 4 | 1.03 [0.94-1.12] | 0 |
|  | 15 mcg | 11 | 1.01[0.99-1.03] | 38.6 |
| Seroprotection B<br>strain | 9 mcg | 4 | 0.95[0.71-1.27] | 0 |
|  | 15 mcg | 12 | 1.03[0.97-1.09] | 45.3 |
| GMT H1N1 | 9 mcg | 4 | 0.96[0.75-1.23] | 99.3 |
|  | 15 mcg | 11 | 1.11 [0.89-1.39] | 100 |
| GMT H3N2 | 9 mcg | 4 | 1.07 [0.80-1.44] | 99.5 |
|  | 15 mcg | 7 | 1.13 [0.92-1.40] | 100 |
| GMT B strain | 9 mcg | 4 | 0.93 [0.72-1.20] | 99.5 |
|  | 15 mcg | 9 | 1.19[1.09-1.30] | 99.8 |

#### 3.3.5 Influenza Infection or Influenza-like Illness

A meta-analysis of four studies reporting clinical outcomes showed that the risk of influenza or influenza-like illness was significantly lower with intradermal vaccines when compared to intramuscular vaccines (RR 0.62, 95% CI: 0.49-0.77). There was, however, no significant difference between the two routes of administration after dose stratification (Figure 2).

**Figure 2.**
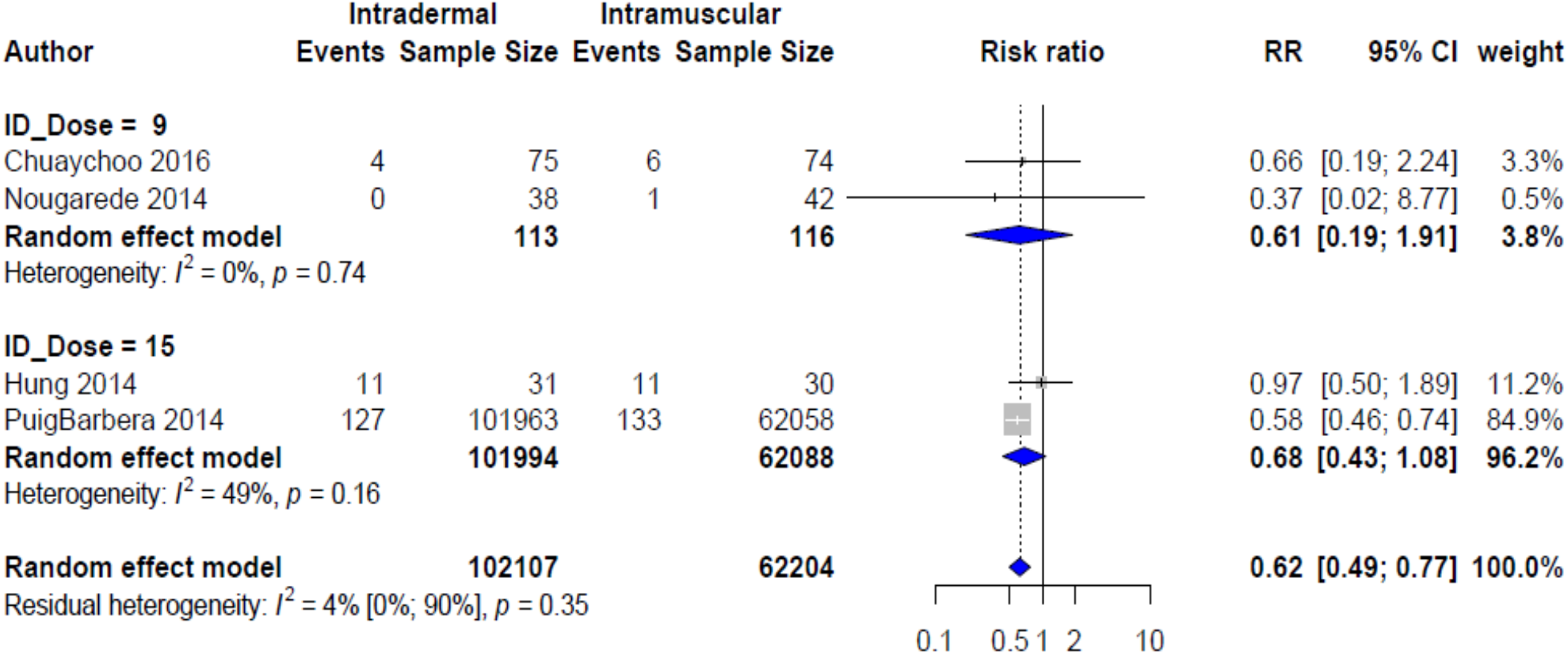
Risk of Influenza and Influenza-like Illness by Route of Vaccine Administration

#### 3.3.6 Adverse Events

Local adverse events, including erythema, swelling, induration, pruritus, and ecchymosis, were significantly higher across the dose spectrum of intradermal vaccines compared with the standard intramuscular dose. However, pain was equivalent between the 6 mcg, 9 mcg, and 15 mcg intradermal doses and the 15 mcg intramuscular dose, but was significantly lower with the 3 mcg intradermal dose (Table 5). Systemic adverse events, including headache, fever, malaise, arthralgia, myalgia, and nausea, were equivalent between the low intradermal doses and the standard intramuscular dose, albeit fever and chills were more common with the 9 mcg intradermal dose (Table 6).

**Table 5.**
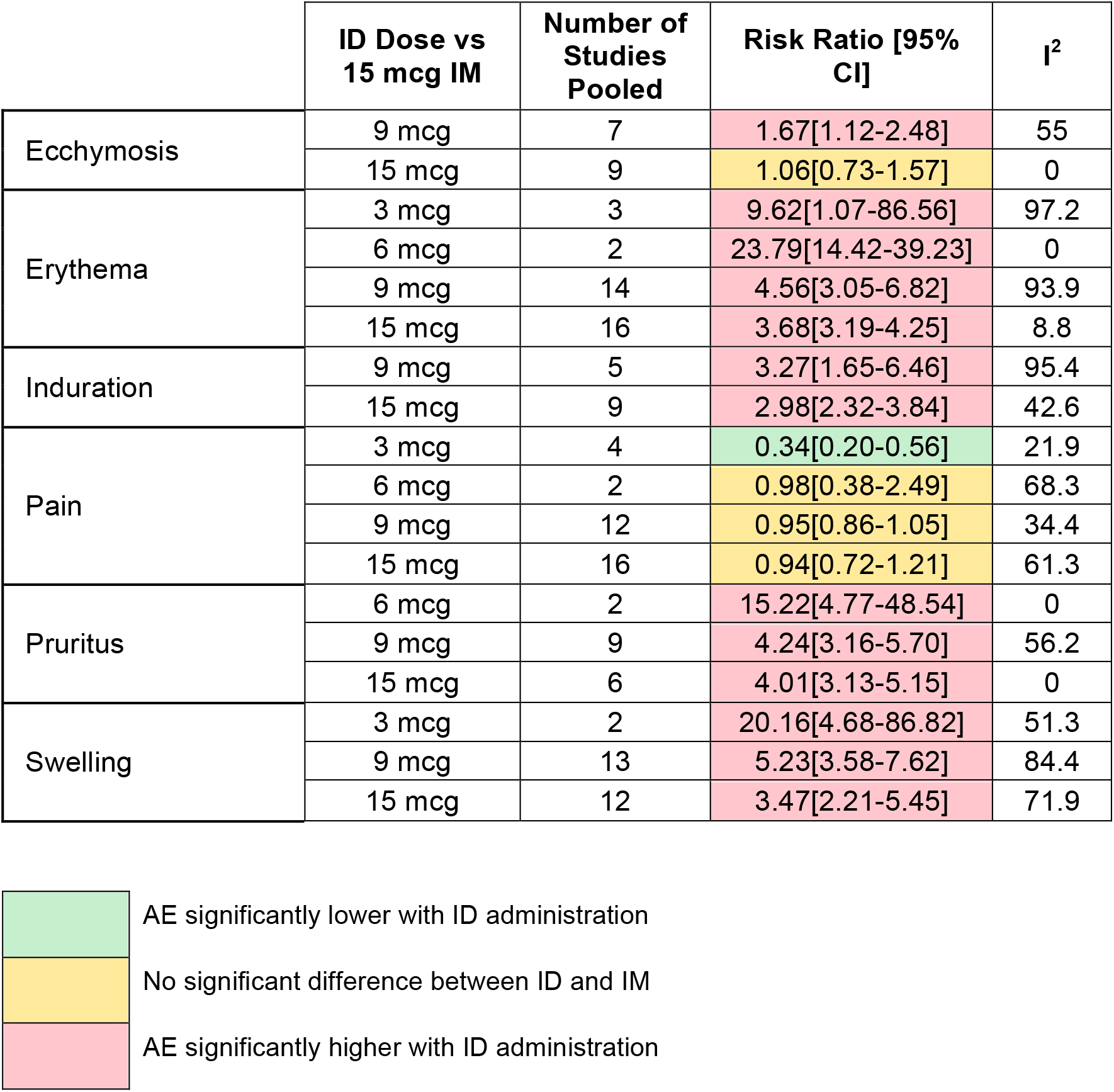
Local Adverse Events Risks with ID and IM Doses

|  | ID Dose vs<br>15 mcg IM | Number of<br>Studies<br>Pooled | Risk Ratio [95%<br>CI] | I<sup>2</sup> |
| --- | --- | --- | --- | --- |
| Ecchymosis | 9 mcg | 7 | 1.67[1.12-2.48] | 55 |
|  | 15 mcg | 9 | 1.06[0.73-1.57] | 0 |
| Erythema | 3 mcg | 3 | 9.62[1.07-86.56] | 97.2 |
|  | 6 mcg | 2 | 23.79[14.42-39.23] | 0 |
|  | 9 mcg | 14 | 4.56[3.05-6.82] | 93.9 |
|  | 15 mcg | 16 | 3.68[3.19-4.25] | 8.8 |
| Induration | 9 mcg | 5 | 3.27[1.65-6.46] | 95.4 |
|  | 15 mcg | 9 | 2.98[2.32-3.84] | 42.6 |
| Pain | 3 mcg | 4 | 0.34[0.20-0.56] | 21.9 |
|  | 6 mcg | 2 | 0.98[0.38-2.49] | 68.3 |
|  | 9 mcg | 12 | 0.95[0.86-1.05] | 34.4 |
|  | 15 mcg | 16 | 0.94[0.72-1.21] | 61.3 |
| Pruritus | 6 mcg | 2 | 15.22[4.77-48.54] | 0 |
|  | 9 mcg | 9 | 4.24[3.16-5.70] | 56.2 |
|  | 15 mcg | 6 | 4.01[3.13-5.15] | 0 |
| Swelling | 3 mcg | 2 | 20.16[4.68-86.82] | 51.3 |
|  | 9 mcg | 13 | 5.23[3.58-7.62] | 84.4 |
|  | 15 mcg | 12 | 3.47[2.21-5.45] | 71.9 |

|  |  |
| --- | --- |
|  | AE significantly lower with ID administration |
|  | No significant difference between ID and IM |
|  | AE significantly higher with ID administration |

**Table 6.** Systemic Adverse Events Risks with ID and IM Doses

|  | ID Dose vs 15 mcg IM | Number of Studies Pooled | Risk Ratio [95% CI] | I<sup>2</sup> |
| --- | --- | --- | --- | --- |
| Arthralgia | 15 mcg | 3 | 1.17[0.39-3.53] | 22.7 |
| Chills and shivering | 9 mcg | 7 | 1.24[1.03-1.50] | 0 |
|  | 15 mcg | 10 | 1.08[0.78-1.51] | 0 |
| Fever | 6 mcg | 2 | 0.54[0.17-1.71] | 34.5 |
|  | 9 mcg | 11 | 1.36[1.03-1.80] | 0 |
|  | 15 mcg | 13 | 0.89[0.59-1.34] | 0 |
| Headache | 3 mcg | 2 | 1.09[0.86-1.37] | 0 |
|  | 6 mcg | 2 | 0.83[0.39-1.78] | 68 |
|  | 9 mcg | 13 | 1.03[0.96-1.11] | 0 |
|  | 15 mcg | 9 | 1.16[0.94-1.45] | 0 |
| Malaise | 9 mcg | 7 | 1.05[0.94-1.20] | 7.1 |
|  | 15 mcg | 14 | 0.97[0.78-1.22] | 0 |
| Myalgia | 9 mcg | 12 | 1.24[0.93-1.65] | 74.8 |
|  | 15 mcg | 9 | 0.84[0.63-1.12] | 29.4 |
| Nausea | 9 mcg | 3 | 0.93[0.37-2.31] | 0 |
|  | 15 mcg | 2 | 1.05[0.33-3.33] | 0 |

#### 3.3.7 Publication Bias

Egger’s test for publication bias was statistically significant for the 15 mcg intradermal and intramuscular comparison for the B and H3N2 strains seroconversion rates (P=0.024, respectively). Bias correction using trim and fill method did not change the statistical significance of the unadjusted results (Figure 3).

**Figure 3.**
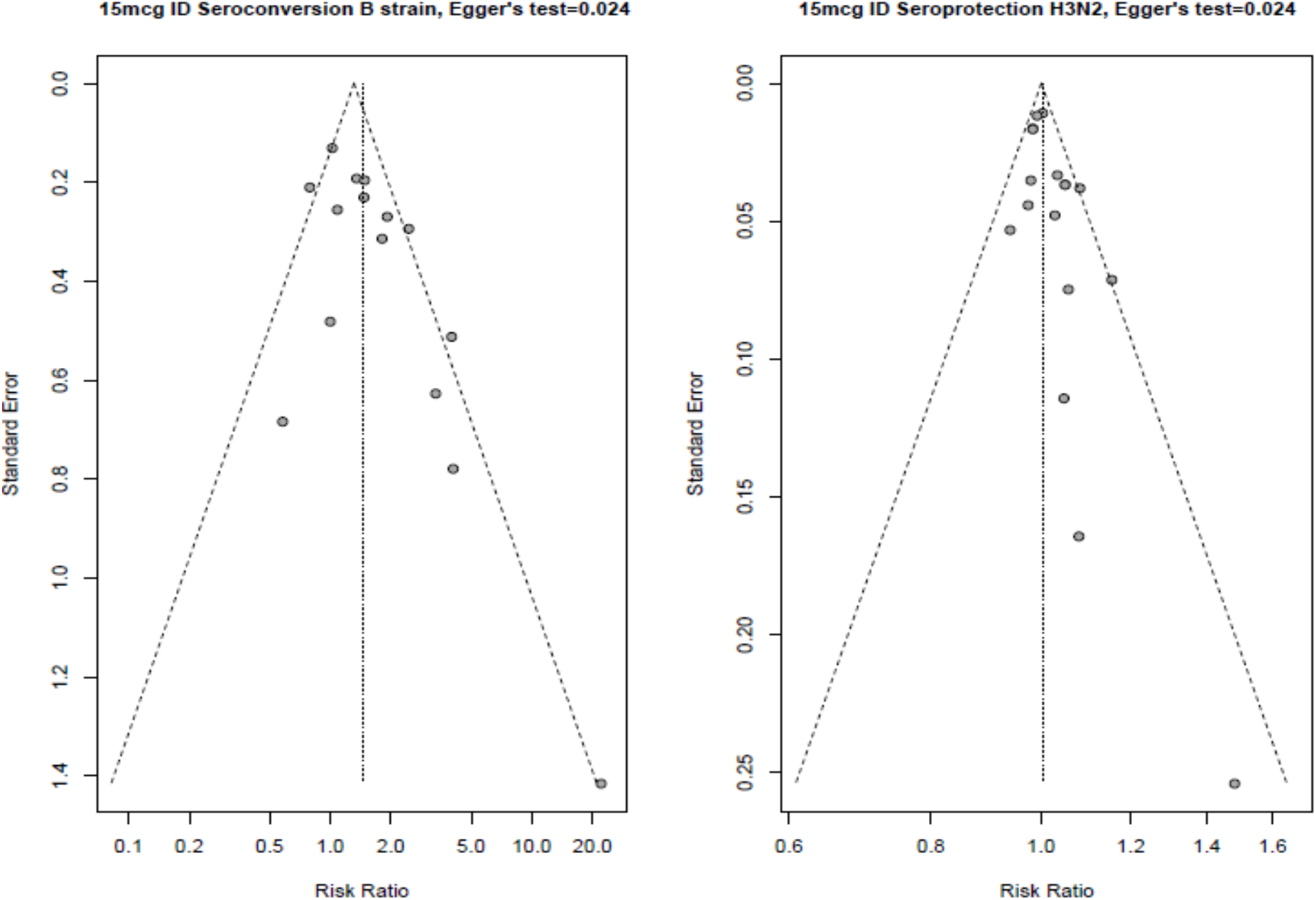
Funnel Plot for Outcomes with Significant Asymmetry

## 4 Discussion

This rapid review and meta-analysis showed that the 9 mcg and 15 mcg intradermal vaccination doses demonstrated immunogenicity that was equivalent to the full-dose 15 mcg intramuscular influenza vaccination, irrespective of patient age. However, the 15 mcg intradermal vaccine showed significantly better immunogenicity for some of the outcomes and strains, suggesting that the immunological response may be dose-related. The risk of local adverse events, such as erythema, induration, swelling, and ecchymosis was reduced with intramuscular vaccination; however, the risk of pain did not differ significantly between the two administration methods, with the exception of the 3 mcg intradermal dose, which significantly lowered the risk of pain. The risks of systemic adverse events, such as headache, malaise, myalgia, and arthralgia, were similar with both administration methods.

The findings of the present study are similar to those by Marra et al.^47^ and Pileggi et al.,^48 49^ which demonstrated equivalence between the different intradermal influenza vaccine doses and the 15 mcg intramuscular influenza vaccine dose. It should be noted that Pileggi et al. included studies involving only immunocompromised participants in one of their studies^48^ and elderly participants in another.^49^ However, the present rapid review excluded immunocompromised patients and carried out sensitivity analysis of studies involving the elderly, given that old age^50^ and immunocompromise^51^ are known to attenuate immunological response. Although local skin reactions were more common with intradermal vaccinations, these reactions are generally well-accepted by vaccinees,^52 53^ who also find the microinjection systems to be more tolerable than the regular needles.^53^

A limitation of this study was the heterogeneity among the included studies, particularly with respect to the GMT outcome. This may be attributed to the variation in the characteristics of the study participants, including age and co-morbidities. However, heterogeneity persisted after stratifying the meta-analyses by age group. Other possible causes of heterogeneity include variations in vaccine factors, such as the use of adjuvants and differences in vaccine brands and delivery systems.

In conclusion, given the similarity in immunogenicity between the low dose intradermal and standard dose intramuscular influenza vaccine, low dose intradermal vaccine may be a reasonable alternative to standard dose intramuscular vaccination.

## Data Availability

All datasets supporting the conclusions of this article are included within the article.

## Acknowledgements

We thank Andrea Tricco and the Drug Safety and Effectiveness Network (DSEN) Methods and Applications Group for Indirect Comparisons (MAGIC) team.

