## Supplemental Files for "Intradermal versus Intramuscular Administration of Influenza Vaccination: Rapid Review and Meta-analysis"

### Supplemental Files: Intradermal versus Intramuscular Administration of Influenza Vaccination

### Appendix A: Search Strategies

**MEDLINE**

1. Influenza, Human/

2. exp influenzavirus a/ or exp influenzavirus b/ or influenzavirus c/

3. Influenza Vaccines/

4. (influenza* or flu).tw,kf.

5. 1 or 2 or 3 or 4

6. Injections, Intradermal/

7. (intradermal or intra-dermal).tw,kf.

8. ID injection*.tw,kf.

9. IDflu.tw,kf.

10. (inject* adj5 (dermal or dermis)).tw,kf.

11. mantoux.tw,kf.

12. dose sparing.tw,kf.

13. 6 or 7 or 8 or 9 or 10 or 11 or 12

14. 5 and 13

15. animals/ not humans/

16. 14 not 15

17. limit 16 to (case reports or editorial or letter)

18. 16 not 17

19. limit 18 to yr="2010 -Current"

**EMBASE**

1. influenza/ or exp influenza a/ or influenza b/ or influenza c/ or seasonal influenza/

2. influenza virus/ or exp influenzavirus a/ or exp influenzavirus b/ or exp influenzavirus c/

3. influenza vaccine/

4. (influenza* or flu).tw,kw.

5. 1 or 2 or 3 or 4

6. intradermal drug administration/

7. (intradermal or intra-dermal).tw,kw.

8. ID injection*.tw,kw.

9. IDflu.tw,kw.

10. (inject* adj5 (dermal or dermis)).tw,kw.

11. mantoux.tw,kw.

12. dose sparing.tw,kw.

13. 6 or 7 or 8 or 9 or 10 or 11 or 12

14. 5 and 13

15. exp influenza vaccine/dl [Intradermal Drug Administration]

16. 14 or 15

17. animals/ not human/

18. 16 not 17

19. limit 18 to (conference abstract or editorial or letter)

20. 18 not 19

21. limit 20 to yr="2010 -Current"

**Cochrane Central Register of Controlled Trials**

1. Influenza, Human/

2. exp influenzavirus a/ or exp influenzavirus b/ or influenzavirus c/

3. Influenza Vaccines/

4. (influenza* or flu).tw.

5. 1 or 2 or 3 or 4

6. Injections, Intradermal/

7. (intradermal or intra-dermal).tw.

8. ID injection*.tw.

9. IDflu.tw.

10. (inject* adj5 (dermal or dermis)).tw.

11. mantoux.tw.

12. dose sparing.tw.

13. 6 or 7 or 8 or 9 or 10 or 11 or 12

14. 5 and 13

15. animals/ not humans/

16. 14 not 15

### Appendix B: Study Characteristics

Table B1. Characteristics of Included Studies

| **Author** | **Inclusion/Exclusion Criteria** | **Demographic** | **Vaccine Information** | **Intervention** | **Analysis** |
| --- | --- | --- | --- | --- | --- |
| **Randomized Controlled Trials** | | | | | |
| **Ansaldi et al.**  Italy  (2013)^18^  Date of Recruitment:  Sept – Dec 2010  Trial Setting:  Multicenter | **Inclusion:**  ≥60 years of age  **Exclusion:**  Systemic hypersensitivity to egg or chicken proteins or any of the vaccine constituents, acute febrile illness (temperature ≥ 37.5°C) at the time of enrolment, any vaccination within the previous 4 weeks or planned vaccination within the 6 months following vaccination, current abuse of alcohol or drug addiction, seasonal influenza vaccination in the previous 6 months with vaccines object of the study or other influenza vaccines and unstable chronic illness that could interfere with the conduction or completion of the study. Subjects with high-risk medical conditions for influenza infection were not excluded, provided they met the above criteria. | **Total Sample Size (% Female):**  n=47 (NR)  **Age Range:**  ≥60 years  **Pre-existing Condition:**  NR | **Vaccine Type:**  Trivalent  **Vaccine Characteristics:**  A/H1N1 (A/California/7/2009,  A/Genoa/1/11, A/Genoa/6/11, A/Genoa/24/11), A/H3N2 (A/Perth/16/2009),  B Strain (B/Brisbane/60/2008)  **Type of Adjuvant:**  Virosome  **Quantity of Adjuvant:**  NR | **Intradermal**  **Sample Size (% Female):**  n=24 (NR)  **Age (Mean ± SD):**  76.9 ± 8.4  **Dose:**  15µg  **Intradermal Administration Technique:**  Microneedle System (Soluvia) | **Analysis Type:**  ITT  **Maximum Follow-up:**  90 days  **Outcomes:**   - Adverse events - GMT - MFI - Seroconversion - Seroprotection |
|  |  |  |  | **Intramuscular**  **Sample Size (% Female):**  n=23 (NR)  **Age (Mean ± SD):**  75.3 ± 7.7  **Dose:**  15µg |  |
| **Ansaldi et al.** France, Belgium, Lithuania, Italy  (2012)  **^17^**  Date of Recruitment:  2006  Trial Setting:  Multicenter | **Inclusion:**  NR  **Exclusion:**  Unstable chronic illness, systemic hypersensitivity, to vaccine constituent, history of life threatening reaction to vaccine or vaccine with same constituent, acute febrile illness, immunodeficiency, immunosuppressive therapy in past 6 months, long term treatment with steroids, alcohol abuse, drug addiction, unstable chronic illness that could interfere with study completion, receipt of blood or blood products in last 3 months, influenza vaccination in previous 6 months, any vaccination within 4 weeks, thrombocytopenia, bleeding disorder | **Total Sample Size (% Female):**  n= 50 (NR)  **Age Range:**  ≥60 years  **Pre-existing Condition:**  NR | **Vaccine Type:**  Trivalent  **Vaccine Characteristics:**  A/H3N2 (Wisconsin/67/05, Genoa62/05,  Genoa03/07,  Brisbane/10/07,  Genoa02/07,  Genoa03/06)  **Type of Adjuvant:**  NR  **Quantity of Adjuvant:**  N/A | **Intradermal**  **Sample Size (% Female):**  n=25 (NR)  **Age (Mean ± SD):**  68.2 ± 7  **Dose:**  15µg  **Intradermal Administration Technique:**  NR | **Analysis Type:**  ITT  **Maximum Follow-up:**  21 days  **Outcomes:**   - GMT - MFI - Seroconversion - Seroprotection |
|  |  |  |  | **Intramuscular**  **Sample Size (% Female):**  n=25 (NR)  **Age (Mean ± SD):**  73.5 ± 7.4  **Dose:**  15µg |  |
| **Arnou et al.** France, Italy, Belgium, Lithuania  (2010)  **^19^**  Date of Recruitment:  Sept – Dec 2006  Trial Setting:  Multicenter | **Inclusion:**  Aged 18 to 60 years. Women were eligible with either a confirmed inability to conceive or a negative urine pregnancy test at the first visit.  **Exclusion:**  Systemic hypersensitivity to egg proteins, chick proteins, or any of the vaccine components; febrile illness on the day of inclusion; congenital or acquired immunodeficiency, immunosuppressive therapy such as anticancer chemotherapy or radiation therapy within the preceding six months or long-term systemic corticosteroids therapy; blood or blood-derived products received in the past three months; any vaccination in the four weeks preceding the trial vaccination or planned vaccination in the four weeks following the trial vaccination; or vaccination against influenza in the previous six months. | **Total Sample Size**  **(% Female):**  n=1676 (59.8)  **Age Range:**  18 – 60 years  **Pre-existing Condition:**  NR | **Vaccine Type:**  Trivalent  **Vaccine Characteristics:**  A/H1N1 (A/New Caledonia/20/99), A/H3N2 (A/Wisconsin/67/2005),  B Strain (B/Malaysia/2506/2004)  **Type of Adjuvant:**  NR  **Quantity of Adjuvant:**  N/A | **Intradermal**  **Sample Size**  **(% Female):**  n=1255 (60.2)  **Age (Mean ± SD):**  NR  **Dose:**  9µg  **Intradermal Administration Technique:**  Microneedle System (Soluvia) | **Analysis Type:**  PP  **Maximum Follow-up:**  21 days  **Outcomes:**   - Adverse events - Serious adverse events - GMT - MFI - Seroconversion - Seroprotection |
|  |  |  |  | **Intramuscular**  **Sample Size**  **(% Female):**  n=421 (59.4)  **Age (Mean ± SD):**  NR  **Dose:**  15 µg |  |
| **Boonnak et al.**  Thailand  (2017)^20^  Date of Recruitment:  2012 – 2013  Trial Setting:  NR | **Inclusion:**  NR  **Exclusion:**  NR | **Total Sample Size (% Female):**  n= 221(80.1)  **Age Range:**  60 – 84 years  **Pre-existing Condition:**  dyslipidemia, Diabetes mellitus, Hypertension | **Vaccine Type:**  Trivalent  **Vaccine Characteristics:**  A/H1N1(A/California/07/09), A/H3N2 (A/Songhka/308/13), B Strain (B/Phuket/287/13)  **Type of Adjuvant:**  NR  **Quantity of Adjuvant:**  N/A | **Intradermal**  **Sample Size (% Female):**  n=111 (NR)  **Age (Mean ± SD):**  66.1 ± 5.1  **Dose:**  15µg | **Analysis Type:**  ITT  **Maximum Follow-up:**  60 days  **Outcomes:**   - Adverse events - GMT - Seroconversion - Seroprotection |
|  |  |  |  | **Intramuscular**  **Sample Size (% Female):**  n=110 (NR)  **Age (Mean ± SD):**  65.5 ± 5.9  **Dose:**  15µg |  |
| **Camiloni et al.**  Italy  (2014)^21^  Date of Recruitment:  2011 – 2012  Trial Setting:  Multicenter | **Inclusion:**  Elderly people living in two nursing homes located in Umbria (Italy) immunized with a single dose of trivalent influenza vaccine in November 2011  **Exclusion:**  NR | **Total Sample Size (% Female):**  n= 80 (85)  **Age Range:**  64 – 100 years  **Pre-existing Condition:**  Cardiovascular disease, Diabetes mellitus, cancer, other chronic conditions | **Vaccine Type:**  Trivalent  **Vaccine Characteristics:**  A/H1N1 (  A/California/7/09,  A/Perugia/06/12, A/Perugia/20/12,  A/Perugia/44/12,  A/Perugia/50/12), A/H3N2 (A/Perth/16/09), B Strain (B/Brisbane/60/08)  **Type of Adjuvant:**  MF59  **Quantity of Adjuvant:**  NR | **Intradermal**  **Sample Size (% Female):**  n=40 (NR)  **Age (Mean ± SD):**  84.6 (NR)  **Dose:**  15µg | **Analysis Type:**  ITT  **Maximum Follow-up:**  180 days  **Outcomes:**   - GMT - MFI - Seroconversion - Seroprotection |
|  |  |  |  | **Intramuscular**  **Sample Size (% Female):**  n=40 (NR)  **Age (Mean ± SD):**  85.3 ± NR  **Dose:**  15µg |  |
| **Carter et al.** USA  (2019)  **^22^**  Date of Recruitment:  Aug 2012 – Jan 2013  Trial Setting:  Multicenter | **Inclusion:**  18 to 70 years old; Available for clinical follow-up through Study Week 40 for Groups 1-2 and through Study Week 64 for Groups 3-6; able and willing to complete the informed consent process; willing to donate blood for sample storage to be used for future research; physical examination and laboratory results without clinically significant findings and a Body Mass Index (BMI) ≤40 within the 70 days prior to enrolment; has not yet received the current year (2012/13) influenza vaccine prior to enrolment and agrees to receive seasonal influenza vaccines during study participation only from the study site; hemoglobin within institutional normal limits; white blood cells either within institutional normal range or accompanied by site physician approval as consistent with healthy adult status; platelets = 125,000 – 500,000/mm3; alanine aminotransferase (ALT) ≤ 2.5 x upper limit of normal (ULN); Serum creatinine ≤ 1 x ULN based on site institutional normal range; Criteria applicable to women of childbearing potential: Negative human chorionic gonadotropin (β-HCG) pregnancy test (urine or serum) on day of enrolment; agree to use an effective means of birth control from 21 days prior to enrolment through 3 weeks after the second study vaccination.  **Exclusion:**  Breast-feeding or planning to become pregnant while participating in the study; subject has received any of the following substances: more than 10 days of systemic immunosuppressive medications or cytotoxic medications within the 12 weeks prior to enrolment or any within the 14 days prior to enrolment, blood products within 16 weeks prior to enrolment; immunoglobulin within 8 weeks prior to enrolment; Investigational research agents within 28 days (4 weeks) prior to enrolment or planning to receive investigational products while on the study; allergy treatment with antigen injections, unless on maintenance schedule and allergy shots could be staggered with the study vaccinations, within 14 days (2 weeks) prior to enrolment; current anti-TB prophylaxis or therapy; subject has a history of any of the following clinically significant conditions: contraindication to receiving an FDA-approved seasonal influenza vaccination; serious reactions to vaccines that preclude receipt of study vaccinations, as determined by the site investigator; hereditary angioedema (HAE), acquired angioedema (AAE), or idiopathic forms of angioedema; asthma that is severe, unstable or required emergent care, urgent care, hospitalization or intubation during the previous two years or that is expected to require the use of oral, intravenous or high dose inhaled corticosteroids; Diabetes mellitus type I; thyroid disease that is not well-controlled; generalized idiopathic urticaria within the 1 year prior to enrolment; hypertension that is not well controlled; bleeding disorder diagnosed by a doctor (e.g. factor deficiency, coagulopathy, or platelet disorder requiring special precautions), or significant bruising or bleeding difficulties with IM injections or blood draws, or use of blood thinners such as Coumadin or Plavix®; malignancy that is active or treated malignancy for which there is not reasonable assurance of sustained cure or malignancy that is likely to recur during the period of the study; seizure disorder other than: 1) febrile seizures, 2) seizures secondary to alcohol withdrawal more than 3 years ago, or 3) seizures for which no treatment has been required within the 3 years prior to enrollment; Asplenia, functional asplenia or any condition resulting in the absence or removal of the spleen; Guillain-Barré Syndrome; psychiatric condition that precludes compliance with the protocol; past or present psychoses; disorder requiring lithium or within 5 years prior to enrollment, a history of suicide plan or attempt; any medical, psychiatric, or other condition that, in the judgment of the investigator, is a contraindication to protocol participation or impairs ability to give informed consent. | **Total Sample Size (% Female):**  n= 106 (63.2)  **Age Range:**  18 – 69 years  **Pre-existing Condition:**  None | **Vaccine Type:**  Trivalent  **Vaccine Characteristics:**  A/H1N1 (A/California/04/2009), A/H3N2 (A/Victoria/361/2011, A/Texas/50/2012), B Strain (B/Texas/6/2011, B/Wisconsin/1/2010,  B/Massachusetts/2/2012)  **Type of Adjuvant:**  NR  **Quantity of Adjuvant:**  N/A | **Intradermal**  **Sample Size (% Female):**  n=49 (NR)  **Age (Mean ± SD):**  41.9 ± 14  **Dose:**  9µg  **Intradermal Administration Technique:**  short-needle microinjection system | **Analysis Type:**  ITT  **Maximum Follow-up:**  21 days  **Outcomes:**   - Adverse events - GMT - Seroconversion |
|  |  |  |  | **Intramuscular**  **Sample Size (% Female):**  n= 56 (NR)  **Age (Mean ± SD):**  41.9 ± 16  **Dose:**  15µg |  |
| **Chan et al.**  Hong Kong  (2014)^23^  Date of Recruitment:  Oct 2013 – Apr 2014  Trial Setting:  Single Center | **Inclusion:**  Nursing home older adults age of 65 or above from one of the 9 nursing homes who satisfied the 3. World Health Organization recommendation for annual vaccination against influenza  **Exclusion:**  Acute febrile illness; hypersensitivity to egg or chicken proteins or any of the vaccine constituents; thrombocytopenia or a bleeding disorder contraindicating intramuscular vaccination;nunstable chronic illness; and dementia. Older adults were also excluded if they had received immunosuppressive therapy within the previous 6 months or received systemic steroid for more than 1 month within the previous 6 months, acute febrile illness; hypersensitivity to egg or chicken proteins or any of the vaccine constituents; thrombocytopenia or a bleeding disorder contraindicating intramuscular vaccination; | **Total Sample Size (% Female):**  n= 100 (64)  **Age Range:**  ≥65 years  **Pre-existing Condition:**  NR | **Vaccine Type:**  Trivalent  **Vaccine Characteristics:**  A/H1N1 (A/Victoria/361/2011), A/H3N2 (A/California/7/2009), B Strain (B/Massachusetts/2/2012)  **Type of Adjuvant:**  NR  **Quantity of Adjuvant:**  N/A | **Intradermal**  **Sample Size (% Female):**  n=50 (NR)  **Age (Mean ± SD):**  NR  **Dose:**  15µg  **Intradermal Administration Technique:**  microinjection system | **Analysis Type:**  ITT  **Maximum Follow-up:**  180 days  **Outcomes:**   - Adverse events - GMT - MFI - Seroconversion - Seroprotection |
|  |  |  |  | **Intramuscular**  **Sample Size (% Female):**  n=50 (NR)  **Age (Mean ± SD):**  NR  **Dose:**  15µg |  |
| **Chi et al.**  USA  (2010)^24^  Date of Recruitment:  Aug 2007 – NR  Trial Setting:  Single Center | **Inclusion:**  Community-dwelling adults age of 65 or older  **Exclusion:**  Serious or unstable conditions such as advanced coronary heart disease or congestive heart failure, oxygen-dependent chronic obstructive lung disease, insulin-requiring diabetes mellitus, cancer (except localized skin or nonmetastatic prostate cancer), and acute or progressive renal, hepatic, or neurologic conditions | **Total Sample Size (% Female):**  n= 130 (17.1)  **Age Range:**  ≥65 years  **Pre-existing Condition:**  Any chronic condition; lung disease; heart disease; Diabetes mellitus, cancer other | **Vaccine Type:**  Trivalent  **Vaccine Characteristics:**  A/H1N1 (A/Solomon Islands/3/2006), A/H3N2 (  A/Wisconsin/67/2005), B Strain (B/Malaysia/2506/2004)  **Type of Adjuvant:**  NR  **Quantity of Adjuvant:**  N/A | **Intradermal**  **Sample Size (% Female):**  n=65 (NR)  **Age (Mean ± SD):**  NR  **Dose:**  9µg  **Intradermal Administration Technique:**  Microinjection system | **Analysis Type:**  ITT  **Maximum Follow-up:**  28 days  **Outcomes:**   - Adverse events - Serious adverse events - GMT - Seroprotection |
|  |  |  |  | **Intramuscular**  **Sample Size (% Female):**  n=65 (NR)  **Age (Mean ± SD):**  NR  **Dose:**  15µg |  |
| **Chuaychoo**  **et al.**  Thailand  (2019)^25^  Date of Recruitment:  2010 – 2011  Trial Setting:  Single Center | **Inclusion:**  Chronic Obstructive Pulmonary Disease (COPD) patients ≥60 years of age, participated in previous study  **Exclusion:**  NR | **Total Sample Size (% Female):**  n= 80 (7.5)  **Age Range:**  ≥60 years  **Pre-existing Condition:**  COPD; patients had other co-morbidities including Hypertension, Diabetes, dyslipidemia, Coronary artery disease, Cerebrovascular accident, previous Tuberculosis, Chronic kidney disease | **Vaccine Type:**  Trivalent  **Vaccine Characteristics:**  A/H1N1 (A/California/7/2009), A/H3N2 (A/Perth/16/2009), B Strain (B/Brisbane/60/2008)  **Type of Adjuvant:**  NR  **Quantity of Adjuvant:**  N/A | **Intradermal**  **Sample Size (% Female):**  n=41 (NR)  **Age (Mean ± SD):**  72.1 ±5.9  **Dose:**  9µg  **Intradermal Administration Technique:**  25G needle: two injections | **Analysis Type:**  ITT  **Maximum Follow-up:**  365 days  **Outcomes:**   - Adverse events - GMT - Seroconversion - Seroprotection |
|  |  |  |  | **Intramuscular**  **Sample Size (% Female):**  n=39 (NR)  **Age (Mean ± SD):**  75.3 ± 9.7  **Dose:**  15µg |  |
| **Chuaychoo**  **et al.**  Thailand  (2016)^26^  Date of Recruitment:  2010 – 2011  Trial Setting:  Single Center | **Inclusion:**  COPD patients who were 60 y or older and had no seasonal influenza vaccination or had a previous seasonal influenza vaccination more than one year prior  **Exclusion:**  Ongoing fever (BT > 38 C), were immunocompromised hosts or receiving any immunosuppressive drugs, including systemic corticosteroids, had malignancy with an expected survival time of less than a year, or an allergy to vaccine components | **Total Sample Size (% Female):**  n= 149 (8.7)  **Age Range:**  60 – 94 years  **Pre-existing Condition:**  COPD; patients had other co-morbidities including Hypertension, Diabetes, dyslipidemia, Coronary artery disease, Cerebrovascular accident | **Vaccine Type:**  Trivalent  **Vaccine Characteristics:**  A/H1N1 (A/California/7/2009), A/H3N2 (A/Perth/16/2009), B Strain (B/Brisbane/60/2008)  **Type of Adjuvant:**  NR  **Quantity of Adjuvant:**  N/A | **Intradermal**  **Sample Size (% Female):**  n=75 (NR)  **Age (Mean ± SD):**  72 ± 8  **Dose:**  9µg  **Intradermal Administration Technique:**  25G needle: two injections | **Analysis Type:**  ITT  **Maximum Follow-up:**  28 days  **Outcomes:**   - Adverse events - GMT - Seroconversion - Seroprotection |
|  |  |  |  | **Intramuscular**  **Sample Size (% Female):**  n=74 (NR)  **Age (Mean ± SD):**  73 ± 7  **Dose:**  15µg |  |
| **Chuaychoo**  **et al.**  Thailand  (2010)^27^  Date of Recruitment:  Jul 2006 – Feb 2007  Trial Setting:  Single Center | **Inclusion:**  COPD patients who had no vaccination or had a previous influenza vaccination more than a year before the enrolment  **Exclusion:**  Ongoing fever, were immunocompromised, immunosuppressive drugs, malignancy, allergy to vaccine component immunocompromised hosts, were undergoing treatment with any immunosuppressive drug, were being treated with systemic corticosteroids, had a malignancy or any disease that would give an expected survival time of less than a year, or a history of an allergy to eggs, chicken protein or vaccine components | **Total Sample Size (% Female):**  n=156 (8)  **Age Range:**  36 – 91 years  **Pre-existing Condition:**  COPD | **Vaccine Type:**  Trivalent  **Vaccine Characteristics:**  A/H1N1 (A/New Caledonia/20/99), A/H3N2  (A/California/7/2004), B Strain (B/Malaysia/2506/2004)  **Type of Adjuvant:**  NR  **Quantity of Adjuvant:**  N/A | **Intradermal**  **Sample Size (% Female):**  n=81 (NR)  **Age (Mean ± SD):**  72 ± 8  **Dose:**  6µg  **Intradermal Administration Technique:**  The half dose (approximately 0.1 ml) of the vaccine was slowly injected into the dermis of the ventral surface of the right forearm. The rest of the vaccine (approximately 0.1 ml) was injected into the left forearm with the same technique (Fig. 1). A pale orange-peel appearance papule immediately appeared which confirmed that the intradermal injection was performed correctly. | **Analysis Type:**  PP  **Maximum Follow-up:**  365 days  **Outcomes:**   - Adverse events - Serious adverse events - GMT - Seroconversion - Seroprotection |
|  |  |  |  | **Intramuscular**  **Sample Size (% Female):**  n=75 (NR)  **Age (Mean ± SD):**  73 ± 7  **Dose:**  15µg |  |
| **Della Cioppa et al.**  Germany, Poland, Belgium  (2014)^28^  Date of Recruitment:  Oct 2008 – Feb 2009  Trial Setting:  Multicenter | **Inclusion:**  Healthy volunteers ≥65 y of age who were mentally competent and in general good health as determined by medical history, a physical examination, and the clinical judgment of the investigators  **Exclusion:**  Immunization with any influenza vaccine within 6mo, any serious dx, hypersensitivity to vaccine components, immunosuppression, drug and alcohol abuse,anticoagulant,fever 6 months before study enrolment; immunization with any experimental influenza vaccine containing adjuvant within 2 y before study enrolment; any serious disease; hypersensitivity to vaccine components; an impaired or altered immune system; known or suspected history of drug or alcohol abuse; history of bleeding diathesis; conditions associated with prolonged bleeding time, or current use of anticoagulation therapy; laboratory-confirmed influenza disease within 12 mo before study enrolment; receipt of another vaccine or investigational agent within 30 d before study enrolment; infection requiring systemic antibiotic or antiviral therapy within 14 d before study enrolment; and fever (oral temperature ≥ 38 °C) within 7 d before study enrolment. | **Total Sample Size (% Female):**  n=257 (52.96)  **Age Range:**  ≥65 years  **Pre-existing Condition:**  None | **Vaccine Type:**  Trivalent  **Vaccine Characteristics:**  A/H3N2 (A/Uruguay/716/2007) | **Intradermal**  **Sample Size (% Female):**  n=47 (51)  **Age (Mean ± SD):**  68.3 ± 3.5  **Dose:**  6µg  **Intradermal Administration Technique:**  MicronJet delivery system from NanoPass Technologies | **Analysis Type:**  PP  **Maximum Follow-up:**  22 days  **Outcomes:**   - Adverse events - GMT - MFI - Seroconversion - Seroprotection |
|  |  |  |  | **Intradermal**  **Sample Size (% Female):**  n=46 (52)  **Age (Mean ± SD):**  69.6 ± 5.1  **Dose:**  12µg  **Intradermal Administration Technique:**  MicronJet delivery system from NanoPass Technologies |  |
|  |  |  |  | **Intramuscular**  **Sample Size (% Female):**  n=44 (60)  **Age (Mean ± SD):**  69.2 ± 3.6  **Dose:**  15µg  **Type of Adjuvant:**  MF59  **Quantity of Adjuvant:**  0% |  |
|  |  |  |  | **Intramuscular**  **Sample Size (% Female):**  n=43 (44)  **Age (Mean ± SD):**  69.2 ± 4.0  **Dose:**  30µg  **Type of Adjuvant:**  MF59  **Quantity of Adjuvant:**  0% |  |
|  |  |  |  | **Intramuscular**  **Sample Size (% Female):**  n=47 (68)  **Age (Mean ± SD):**  68.2 ± 3.1  **Dose:**  15µg  **Type of Adjuvant:**  MF59  **Quantity of Adjuvant:**  100 |  |
|  |  |  |  | **Intramuscular**  **Sample Size (% Female):**  n=43 (54)  **Age (Mean ± SD):**  69.0 ± 3.5  **Dose:**  30µg  **Type of Adjuvant:**  MF59  **Quantity of Adjuvant:**  100% |  |
| **Esposito et al.**  Italy  (2011)^29^  Date of Recruitment:  Oct – Dec 2010  Trial Setting:  Multicenter | **Inclusion:**  Aged 3 or more years admitted to the clinic for a control examination after a previous hospitalisation for minor surgical problems and who had previously received an influenza vaccination  **Exclusion:**  Chronic disease, a known allergy to any vaccine component, and acute respiratory disease in past 30 days any acute infectious or respiratory disease requiring systemic treatment in the 30 days preceding the start of the study | **Total Sample Size (% Female):**  n= 112 (52)  **Age Range:**  ≥3 years  **Pre-existing Condition:**  NR | **Vaccine Type:**  Trivalent  **Vaccine Characteristics:**  A/H1N1 (A/California/7/2009), A/H3N2 (A/Perth/16/2009), B Strain (B/Brisbane/60/2008)  **Type of Adjuvant:**  Virosome  **Quantity of Adjuvant:**  N/A | **Intradermal**  **Sample Size (% Female):**  n=38 (NR)  **Age (Mean ± SD):**  6.01 ± 2.99  **Dose:**  9µg  **Intradermal Administration Technique:**  microinjection system (Soluvia) | **Analysis Type:**  ITT  **Maximum Follow-up:**  28 days  **Outcomes:**   - Adverse events - GMT - MFI - Seroconversion - Seroprotection |
|  |  |  |  | **Intradermal**  **Sample Size (% Female):**  n=37 (NR)  **Age (Mean ± SD):**  6.11 ± 2.88  **Dose:**  15µg  **Intradermal Administration Technique:**  Microinjection system (Soluvia) |  |
|  |  |  |  | **Intramuscular**  **Sample Size (% Female):**  n=37 (NR)  **Age (Mean ± SD):**  6.1 ± 2.76  **Dose:**  15µg |  |
| **Frenck et al.**  USA  (2011)^30^  Date of Recruitment:  Nov – Dec 2005  Trial Setting:  Multicenter | **Inclusion:**  Previously healthy adults 18–64 years of age  **Exclusion:**  History of immunodeficiency, receipt of blood or blood products within the last 3 months, influenza vaccine within the last 6 months, allergy to components of the vaccine or a condition which, in the opinion of the investigator, may make it unsafe to enrol in the study | **Total Sample Size (% Female):**  n= 1571 (67)  **Age Range:**  18 – 64 years  **Pre-existing Condition:**  None | **Vaccine Type:**  Trivalent  **Vaccine Characteristics:**  A/H1N1 (A/New Caledonia/20/99 IVR-116) , A/H3N2 (A/Wyoming/03/2003 (an A/Fujian/411/2002-like strain)  ), B Strain (B/Jiangsu/10/2003 (a B/Jiangsu/361/2002-like strain))  **Type of Adjuvant:**  NR  **Quantity of Adjuvant:**  N/A | **Intradermal**  **Sample Size (% Female):**  n=395 (NR)  **Age (Mean ± SD):**  NR  **Dose:**  3µg  **Intradermal Administration Technique:**  novel microneedle delivery system (BD Soluvia Microinjection System | **Analysis Type:**  PP  **Maximum Follow-up:**  21 days  **Outcomes:**   - Adverse events - Serious adverse events - GMT - MFI - Seroprotection |
|  |  |  |  | **Intradermal**  **Sample Size (% Female):**  n=390 (NR)  **Age (Mean ± SD):**  NR  **Dose:**  6µg  **Intradermal Administration Technique:**  novel microneedle delivery system (BD Soluvia Microinjection System |  |
|  |  |  |  | **Intradermal**  **Sample Size (% Female):**  n=392 (NR)  **Age (Mean ± SD):**  NR  **Dose:**  9µg  **Intradermal Administration Technique:**  Novel microneedle delivery system (BD Soluvia Microinjection System |  |
|  |  |  |  | **Intramuscular**  **Sample Size (% Female):**  n=394 (NR)  **Age (Mean ± SD):**  NR  **Dose:**  15µg |  |
| **Garg et al.**  Thailand  (2016)^31^  Date of Recruitment:  2011 – 2013  Trial Setting:  Single Center | **Inclusion:**  Thai nationality, Men having Sex with men and 18–60 years old.  **Exclusion:**  Females, men >60 years of age, non-Thai nationality, severe allergies to chicken eggs, prior severe reaction to influenza vaccine, history of Guillain-Barré syndrome, on steroid therapy or other immunosuppressant medications, receipt of influenza vaccine in the 12 months prior to enrollment, receipt of any vaccine in the 4 weeks prior to the first study visit or anticipated receipt of a vaccine (other than influenza vaccine provided through the study protocol) in the 4 weeks following the first study visit, receipt of any experimental agents within the 4 weeks prior to enrollment or anticipated receipt of an experimental agent during the 12 month study period, and any condition, in the opinion of the investigators, that would place them at unacceptable risk of injury or render them unable to meet requirements of the protocol. | **Total Sample Size (% Female):**  n= 80 (0)  **Age Range:**  18 – 60 years  **Pre-existing Condition:**  NR | **Vaccine Type:**  Trivalent  **Vaccine Characteristics:**  A/H1N1, A/H3N2, B Strain  **Type of Adjuvant:**  NR  **Quantity of Adjuvant:**  N/A | **Intradermal**  **Sample Size (% Female):**  n=40 (NR)  **Age (Mean ± SD):**  29 ± NR  **Dose:**  15µg  **Intradermal Administration Technique:**  NR | **Analysis Type:**  ITT  **Maximum Follow-up:**  30 days  **Outcomes:**   - Adverse events - GMT - MFI - Seroconversion - Seroprotection |
|  |  |  |  | **Intramuscular**  **Sample Size (% Female):**  n=40 (NR)  **Age (Mean ± SD):**  30 ± NR  **Dose:**  15µg |  |
| **Gorse et al.**  USA  (2013)^32^  Date of Recruitment:  Oct 2008  Trial Setting:  Multicenter | **Inclusion:**  18–64 years of age and medically stable such that if chronic illness were present it would not interfere with trial conduct or completion  **Exclusion:**  Pregnancy, Gullian barre syndrome, allergy to vaccine components, immunodefficiency, receipt of blood products, febrile/acute illness on day of vaccination immunodeficiency, receipt of blood products in the previous three months, bleeding disorder, history of Guillain-Barré Syndrome, allergy to vaccine components, significant alcohol or drug use, any vaccination in the previous four weeks, vaccination against influenza in the previous six months, and febrile or acute illness on the day of vaccination | **Total Sample Size (% Female):**  n= 3868 (63)  **Age Range:**  18 – 64 years  **Pre-existing Condition:**  None | **Vaccine Type:**  Trivalent  **Vaccine Characteristics:**  A/H1N1 (A/Brisbane/59/07), A/H3N2 (A/Uruguay/716/2007 X-175CA), B Strain (B/Florida/04/2006 Yamagata-like)  **Type of Adjuvant:**  NR  **Quantity of Adjuvant:**  N/A | **Intradermal**  **Sample Size (% Female):**  n=2581 (NR)  **Age (Mean ± SD):**  42.6 ± 13.5  **Dose:**  9µg  **Intradermal Administration Technique:**  1.5 mm length, 30 gauge needle (BD SoluviaTM Microinjection System | **Analysis Type:**  PP  **Follow-up:**  28 days  **Outcomes:**   - Adverse events - Serious adverse events - GMT - Seroconversion - Seroprotection |
|  |  |  |  | **Intramuscular**  **Sample Size (% Female):**  n=1287 (NR)  **Age (Mean ± SD):**  42.5 ± 13.55  **Dose:**  15µg |  |
| **Han et al.**  South Korea  (2013)^33^  Date of Recruitment:  Oct 2010 – Feb 2011  Trial Setting:  Multicenter | **Inclusion:**  ≥18y of age; women of childbearing potential had to be using an effective method of contraception or to abstain from sexual activity for the four weeks prior to vaccination through the four weeks after vaccination  **Exclusion:**  Systemic hypersensitivity to eggs, chicken proteins, or any of the vaccine components; a history of a life-threatening reaction to any vaccine used in the trial or to a vaccine containing any of the same substances; pregnant or had a positive urine pregnancy test; currently breastfeeding a child; received either a seasonal or pandemic influenza vaccine within the previous 12 mo or any other vaccine within the four weeks preceding the trial; an underlying chronic disease including end stage renal disease requiring dialysis, chronic liver disease, active neoplastic disease or hematologic malignancy, received immunosuppressive therapy within the preceding 6 months; received long-term systemic corticosteroid therapy; a contraindication for IM or other vaccination, including thrombocytopenia or anticoagulant therapy; seropositive for human immunodeficiency virus, hepatitis B or hepatitis C; received blood or blood-derived products that might interfere with assessment of the immune response within the previous 3 months | **Total Sample Size (% Female):**  n= 120 (68)  **Age Range:**  ≥18 years  **Pre-existing Condition:**  None | **Vaccine Type:**  Trivalent  **Vaccine Characteristics:**  A/H1N1 (A/California/7/2009), A/H3N2 (A/Perth/16/2009), B Strain (B/Brisbane/60/2008)  **Type of Adjuvant:**  NR  **Quantity of Adjuvant:**  N/A | **Intradermal**  **Sample Size (% Female):**  n=60 (NR)  **Age (Mean ± SD):**  33.0 ± 9.2  **Dose:**  9µg  **Intradermal Administration Technique:**  Soluvia™ microinjection device | **Analysis Type:**  ITT  **Maximum Follow-up:**  21 days  **Outcomes:**   - Adverse events - GMT - Seroconversion - Seroprotection |
|  |  |  |  | **Intradermal**  **Sample Size (% Female):**  n=60 (NR)  **Age (Mean ± SD):**  34.5 ± 10.2  **Dose:**  15µg  **Intradermal Administration Technique:**  Soluvia™ microinjection device |  |
|  |  |  |  | **Intramuscular**  **Sample Size (% Female):**  n=60 (NR)  **Age (Mean ± SD):**  64.5 ± 3.8  **Dose:**  15µg |  |
| **Hung et al.** China  (2016)  **^34^**  Date of Recruitment:  Mar – May 2014  Trial Setting:  Single Center | **Inclusion:**  No influenza vaccine in previous 3 months  **Exclusion:**  History of allergy to vaccine components. | **Total Sample Size (% Female):**  n= 160 (50)  **Age Range:**  18 – 30 years  **Pre-existing Condition:**  NR | **Vaccine Type:**  Trivalent  **Vaccine Characteristics:**  A/H1N1 (A/California/07/2009,  Prototype A/WSN/1933,  A/HK/408027/09, A/H3N2 (A/Victoria/361/2011,  A/HK/485197/14), B Strain (B/Massachusetts/2/2012), Others (B/HK/418078/11)  **Type of Adjuvant:**  NR  **Quantity of Adjuvant:**  N/A | **Intradermal**  **Sample Size (% Female):**  n=80 (NR)  **Median Age (IQR):**  20 (19-22)  **Dose:**  15µg  **Intradermal Administration Technique:**  Soluvia | **Analysis Type:**  ITT  **Maximum Follow-up:**  21 days  **Outcomes:**   - Adverse events - GMT - MFI - Seroconversion - Seroprotection |
|  |  |  |  | **Intramuscular**  **Sample Size (% Female):**  n=80 (NR)  **Median Age (IQR):**  19 (19-21)  **Dose:**  15µg |  |
| **Hung et al.**  China  (2014)^36^  Date of Recruitment:  Jan – Mar 2012  Trial Setting:  Multicenter | **Inclusion:**  All adult patients age ≥ 21 years, attending the medical specialist outpatient clinic in the Hong Kong West Cluster Hospitals, All patients give written informed consent, Subjects must be available to complete the study and comply with study procedures, Willingness to allow for serum samples to be stored beyond the study period, for potential additional future testing to better characterize immune response.  **Exclusion:**  Clinically significant immune-related diseases or significant recent co-morbidities, Inability to comprehend and to follow all required study procedures, History or any illness that might interfere with the results of the study or pose additional risk to the subjects due to participation in the study, Have already received 2011/2012 trivalent influenza vaccine, Have a recent history (documented, confirmed or suspected) of a flu-like disease within a week of vaccination, Have a known allergy to eggs or other components of the study vaccine (including gelatin, formaldehyde, octoxinol, thimerosal, and chicken protein), or history of any anaphylaxis, serious vaccine reactions, to any excipients, Have a positive urine or serum pregnancy test within 24 hours prior to vaccination, or women who are breastfeeding, Female of childbearing potential, not using any acceptable contraceptive methods for at least 2 months prior to study entry or that do not plan to use acceptable birth control measures during the first 3 weeks after vaccination, Have immunosuppression as a result of an underlying illness or treatment, or use of anticancer chemotherapy or radiation therapy (cytotoxic) within the preceding 36 months, Have an active neoplastic disease or a history of any hematologic malignancy, Have long-term use of glucocorticoids including oral, parenteral or high-dose inhaled steroids (>800 mcg/day of beclomethasone dipropionate or equivalent) within the preceding 6 months. (Nasal and topical steroids are allowed), Have a history of receiving immunoglobulin or other blood product within the 3 months prior to vaccination in this study, Have known active human immunodeficiency virus (HIV), hepatitis B, hepatitis C infection or autoimmune hepatitis and related cirrhosis., Received an experimental agent (vaccine, drug, biologic, device, blood product, or medication) within 1 month prior to vaccination in this study or expect to receive an experimental agent during this study, Unwilling to refuse participation in another clinical study through the end of this study, History of progressive or severe neurological disorders, Have received any licensed vaccines within 4 weeks prior to vaccination in this study or plan receipt of such vaccines within 21 days, Axillary temperature ≥ 38°C or oral temperature ≥ 38.5°C within 3 days of intended study vaccination, Surgery planned during the study period that in the Investigator's opinion would interfere with the study visits schedule, Have a history of alcohol or drug abuse in the last 5 years, Have a history of Guillain-Barré Syndrome, Have any condition that the investigator believes may interfere with successful completion of the study. | **Total Sample Size (% Female):**  n= 93 (42.9)  **Age Range:**  ≥21 years  **Pre-existing Condition:**  Hypertension. Diabetes mellitus, Cancer, Chronic renal failure, COPD, Rheumatoid arthritis, Gout, Cerebrovascular accident | **Vaccine Type:**  Trivalent  **Vaccine Characteristics:**  A/H1N1 (A/California/07/2009), A/H3N2 (A/Perth/16/2009), B Strain (B/Brisbane/60/2008)  **Type of Adjuvant:**  NR  **Quantity of Adjuvant:**  N/A | **Intradermal**  **Sample Size (% Female):**  n=31 (NR)  **Median Age (IQR):**  70 (64-79)  **Dose:**  15µg  **Intradermal Administration Technique:**  Soluvia | **Analysis Type:**  PP  **Maximum Follow-up:**  365 days  **Outcomes:**   - Adverse events - GMT - MFI - Seroconversion - Seroprotection |
|  |  |  |  | **Intramuscular**  **Sample Size (% Female):**  n=31 (NR)  **Median Age (IQR):**  75 (60-82)  **Dose:**  15µg |  |
| **Hung et al.**  China  (2012)^35^  Date of Recruitment:  Nov 2010 – Feb 2011  Trial Setting:  Single Center | **Inclusion:**  NR  **Exclusion:**  Clinically, immune dx, recent comorbidity, history of allergy to vaccine component | **Total Sample Size (% Female):**  n= 262 (33.17)  **Age Range:**  ≥21 years  **Pre-existing Condition:**  Chronic diseases (hypertension, diabetes, ischemic heart disease, cancer, stroke, COPD, atrial fibrillation, chronic renal failure) | **Vaccine Type:**  Trivalent  **Vaccine Characteristics:**  A/H1N1 (A/California/07/2009), A/H3N2 (A/Perth/16/2009), B Strain (B/Brisbane/60/2008)  **Type of Adjuvant:**  NR  **Quantity of Adjuvant:**  N/A | **Intradermal**  **Sample Size (% Female):**  n=63 (NR)  **Median Age (IQR):**  72 (68-77)  **Dose:**  3µg  **Intradermal Administration Technique:**  Microneedle | **Analysis Type:**  PP  **Maximum Follow-up:**  21 days  **Outcomes:**   - Adverse events - GMT - MFI - Seroconversion - Seroprotection |
|  |  |  |  | **Intradermal**  **Sample Size (% Female):**  n=68 (NR)  **Median Age (IQR):**  73.5 (69.3-78)  **Dose:**  9µg  **Intradermal Administration Technique:**  Soluvia device |  |
|  |  |  |  | **Intramuscular**  **Sample Size (% Female):**  n=66 (NR)  **Median Age (IQR):**  72 (66-78)  **Dose:**  15µg |  |
| **Leung et al.**  USA  (2017)^37^  Date of Recruitment:  NR  Trial Setting:  Multicenter | **Inclusion:**  Male and female subjects 18 to 64 years of age, inclusive, on the day of vaccination, Had active mild-to-severe Atopic dermatitis (lesions present) with or without a  history of eczema herpeticum or were nonatopic, as diagnosed by using  the ADRN Standard Diagnostic Criteria  **Exclusion:**  Pregnant, allergy to vaccine component, congenital or acquired immunity, systemic steroid for 2 or more weeks, cumulative dose of intranasal or ingled steroid, chronic illness, neoplastic disease or any haematologic malignancy, skin disease other than atopic dermatitis, previous vaccination against influenza in past 6 months, any other live vaccine within 4 weeks, thrombocytopaenia, history of Gullian-barre, phototherapy within 5 days of phototherapy. | **Total Sample Size (% Female):**  n= 336 (55.95)  **Age Range:**  18 – 64 years  **Pre-existing Condition:**  atopic dermatitis | **Vaccine Type:**  Trivalent  **Vaccine Characteristics:**  A/H1N1 (A/California/07/2009), A/H3N2 (A/Perth/16/2009), B Strain (B/Brisbane/60/2008)  **Type of Adjuvant:**  NR  **Quantity of Adjuvant:**  N/A | **Intradermal**  **Sample Size (% Female):**  n=100 (56)  **Age (Mean ± SD):**  35.4 **±** 11.3  **Dose:**  NR  **Intradermal Administration Technique:**  NR | **Analysis Type:**  PP  **Follow-up:**  28 days  **Outcomes:**   - Adverse events - Serious adverse events - GMT - MFI - Seroconversion - Seroprotection |
|  |  |  |  | **Intramuscular**  **Sample Size (% Female):**  n=102 (55.9)  **Age (Mean ± SD):**  36.6 ± 12.1  **Dose:**  NR |  |
| **Levin et al.**  NR  (2016)^45^  Date of Recruitment:  NR  Trial Setting:  NR | **Inclusion:**  Medically stable, healthy participants (P65 years) who were vaccinated against influenza in season 2011–2012,  **Exclusion:**  Previous influenza 2011-2012 vaccination, history of SAE,previous allergy to influenza vaccine, acute excercerbation of bronchopulmonary infection, acute febrile illness | **Total Sample Size (% Female):**  n= 370 (46)  **Age Range:**  ≥65 years  **Pre-existing Condition:**  NR | **Vaccine Type:**  Trivalent  **Vaccine Characteristics:**  A/H1N1 (A/California/07/2009), A/H3N2 (A/Victoria/361/2011), B Strain (B/Wisconsin/1/2010)  **Type of Adjuvant:**  Virosome  **Quantity of Adjuvant:**  N/A | **Intradermal**  **Sample Size (% Female):**  n=61 (NR)  **Age (Mean ± SD):**  NR  **Dose:**  7.5µg  **Intradermal Administration Technique:**  Microneedle | **Analysis Type:**  ITT  **Maximum Follow-up:**  90 days  **Outcomes:**   - Adverse events - Seroconversion - Seroprotection |
|  |  |  |  | **Intradermal**  **Sample Size (% Female):**  n=61 (NR)  **Age (Mean ± SD):**  NR  **Dose:**  15µg  **Intradermal Administration Technique:**  Microneedle |  |
|  |  |  |  | **Intramuscular**  **Sample Size (% Female):**  n=63 (NR)  **Age (Mean ± SD):**  NR  **Dose:**  15µg |  |
| **Levin et al.**  Switzerland  (2014)^44^  Date of Recruitment:  Sept – Nov 2007  Trial Setting:  NR | **Inclusion:**  NR  **Exclusion:**  NR | **Total Sample Size (% Female):**  n= 280 (NR)  **Age Range:**  18 – 60 years  **Pre-existing Condition:**  NR | **Vaccine Type:**  Trivalent  **Vaccine Characteristics:**  A/H1N1 (A/SolomonIslands/3/2006), A/H3N2 (A/Wisconsin/67/2005), B Strain (B/Malaysia/2506/2004)  **Type of Adjuvant:**  Virosome  **Quantity of Adjuvant:**  N/A | **Intradermal**  **Sample Size (% Female):**  n=55 (NR)  **Age (Mean ± SD):**  NR  **Dose:**  3µg  **Intradermal Administration Technique:**  Microneedle | **Analysis Type:**  PP  **Maximum Follow-up:**  21 days  **Outcomes:**   - Adverse events - GMT - MFI - Seroconversion - Seroprotection |
|  |  |  |  | **Intradermal**  **Sample Size (% Female):**  n=53 (NR)  **Age (Mean ± SD):**  NR  **Dose:**  4.5µg  **Intradermal Administration Technique:**  Regular needle |  |
|  |  |  |  | **Intradermal**  **Sample Size (% Female):**  n=55 (NR)  **Age (Mean ± SD):**  NR  **Dose:**  6µg  **Intradermal Administration Technique:**  Regular needle |  |
|  |  |  |  | **Intramuscular**  **Sample Size (% Female):**  n=54 (NR)  **Age (Mean ± SD):**  NR  **Dose:**  15µg |  |
| **Nougarede et al.**  France  (2014)^38^  Date of Recruitment:  Sept 2007 – May 2008  Trial Setting:  Single Center | **Inclusion:**  Healthy adults 18 to 40 y old  **Exclusion:**  Immunodeficiency, hypersensitivity, immunosuppressant, blood transfusion in past 3 months, previous influenza vaccine <6mo,hepatitis B or C, pregnancy, febrile illness or severe acute illness or infection on day of vaccination | **Total Sample Size (% Female):**  n= 80 (69.9)  **Age Range:**  18 – 40 years  **Pre-existing Condition:**  NR | **Vaccine Type:**  Trivalent  **Vaccine Characteristics:**  A/H1N1 (A/SolomonIslands/3/2006), A/H3N2 (A/Wisconsin/67/2005), B Strain (B/Malaysia/2506/2004)  **Type of Adjuvant:**  NR  **Quantity of Adjuvant:**  N/A | **Intradermal**  **Sample Size (% Female):**  n=38 (68.4)  **Age (Mean ± SD):**  26.6 ±5.7  **Dose:**  9µg  **Intradermal Administration Technique:**  Soluvia | **Analysis Type:**  ITT  **Maximum Follow-up:**  180 days  **Outcomes:**   - Adverse events - GMT - MFI - Seroconversion - Seroprotection |
|  |  |  |  | **Intramuscular**  **Sample Size (% Female):**  n=42 (71.4)  **Age (Mean ± SD):**  25.7 ± 4.8  **Dose:**  15µg |  |
| **Patel et al.**  USA  (2010)^39^  Date of Recruitment:  Jul 2005 - NR  Trial Setting:  Single Center | **Inclusion:**  NR  **Exclusion:**  NR | **Total Sample Size (% Female):**  n= 100 (50)  **Age Range:**  18 – 40 years  **Pre-existing Condition:**  NR | **Vaccine Type:**  Not Trivalent  **Vaccine Characteristics:**  H5N1 (A/Vietnam/1203/2004)  **Type of Adjuvant:**  NR  **Quantity of Adjuvant:**  N/A | **Intradermal**  **Sample Size (% Female):**  n=25 (NR)  **Age (Mean ± SD):**  NR  **Dose:**  3µg  **Intradermal Administration Technique:**  Regular needle | **Analysis Type:**  ITT  **Maximum Follow-up:**  28 days  **Outcomes:**   - Adverse events - GMT - Seroconversion - Seroprotection |
|  |  |  |  | **Intradermal**  **Sample Size (% Female):**  n=25 (NR)  **Age (Mean ± SD):**  NR  **Dose:**  9µg  **Intradermal Administration Technique:**  Regular needle |  |
|  |  |  |  | **Intramuscular**  **Sample Size (% Female):**  n=25 (NR)  **Age (Mean ± SD):**  NR  **Dose:**  15µg |  |
|  |  |  |  | **Intramuscular**  **Sample Size (% Female):**  n=25 (NR)  **Age (Mean ± SD):**  NR  **Dose:**  45µg |  |
| **Seo et al.**  South Korea  (2014)^40^  Date of Recruitment:  Oct 2011 – NR  Trial Setting:  Multicenter | **Inclusion:**  Aged 65 years who were not vaccinated with influenza vaccine during the 2011-2012 season and had not been previously diagnosed with influenza infection were recruited at two centers during the first week of Oct 2011  **Exclusion:**  Egg allergy, febrile illness (a temperature of ≥37.5°C) on the day of vaccination, influenza vaccination within the previous 6 months, any other vaccinations within the previous 30 days, high-dose systemic steroid therapy (i.e., ≥0.5 mg/kg of body weight prednisone daily) in the previous 30 days, treatment with immunoglobulins during the previous 3 months, development of influenza-like illness during the vaccination study period, and any conditions that might interfere with the study results. | **Total Sample Size (% Female):**  n= 354 (65.4)  **Age Range:**  ≥65 years  **Pre-existing Condition:**  NR | **Vaccine Type:**  Trivalent  **Vaccine Characteristics:**  A/H1N1 (A/California/7/2009), A/H3N2 (A/Perth/16/2009), B Strain (B/Brisbane/60/2008)  **Type of Adjuvant:**  MF59  **Quantity of Adjuvant:**  N/A | **Intradermal**  **Sample Size (% Female):**  n=111 (NR)  **Mean Age (Range):**  72 (65-86)  **Dose:**  15µg  **Intradermal Administration Technique:**  NR | **Analysis Type:**  PP  **Maximum Follow-up:**  180 days  **Outcomes:**   - Adverse events - GMT - MFI - Seroconversion - Seroprotection |
|  |  |  |  | **Intramuscular**  **Sample Size (% Female):**  n=113 (NR)  **Mean Age (Range):**  73 (65-88)  **Dose:**  15µg |  |
| **Song et al.**  South Korea  (2013)^41^  Date of Recruitment:  2006 - 2007  Trial Setting:  NR | **Inclusion:**  Healthy, 18-30 years old  **Exclusion:**  Immunosuppressant, hypersensitivity to vaccine component, Hx of Gullian Barre, thrombocytopenia. Coagulopathy, febrile illness | **Total Sample Size (% Female):**  n= 96 (NR)  **Age Range:**  18 – 30 years  **Pre-existing Condition:**  NR | **Vaccine Type:**  Trivalent  **Vaccine Characteristics:**  A/H1N1 (A/New Caledonia/20/99), A/H3N2 (  A/Wisconsin/67/2005), B Strain (B/Malaysia/2506/2004)  **Type of Adjuvant:**  NR  **Quantity of Adjuvant:**  N/A | **Intradermal**  **Sample Size (% Female):**  n=30 (NR)  **Age (Mean ± SD):**  24.7 ± 1.6  **Dose:**  3µg  **Intradermal Administration Technique:**  NR | **Analysis Type:**  PP  **Maximum Follow-up:**  180 days  **Outcomes:**   - GMT - Seroconversion - Seroprotection |
|  |  |  |  | **Intradermal**  **Sample Size (% Female):**  n=30 (NR)  **Age (Mean ± SD):**  25.3 ± 2.1  **Dose:**  7.5µg  **Intradermal Administration Technique:**  NR |  |
|  |  |  |  | **Intramuscular**  **Sample Size (% Female):**  n=32 (NR)  **Age (Mean ± SD):**  25.3 ± 2.3  **Dose:**  15µg |  |
| **Tsang et al.**  USA  (2014)^42^  Date of Recruitment:  Oct 2007 – Jun 2008  Trial Setting:  Multicenter | **Inclusion:**  Medically stable, ambulatory, older adults (≥65 years of age) or younger adults (18–49 years of age). Women could not be pregnant or breastfeeding and if of child-bearing potential had to be using an effective method of contraception within 4 weeks before and after vaccination  **Exclusion:**  Non pregnant and not breast feeding, sensitivity to vaccine component, vaccinated for influenza in last 6 months, long term systemic steroids, bleeding disorder, anticoagulant, hepatitis B and C, blood transfusion in last 3 months | **Total Sample Size (% Female):**  n= 1912 (NR)  **Age Range:**  ≥65 years  **Pre-existing Condition:**  NR | **Vaccine Type:**  Trivalent  **Vaccine Characteristics:**  A/H1N1 (A/Solomon Islands/3/2006), A/H3N2 (  A/Wisconsin/67/2005), B Strain (B/Malaysia/2506/2004)  **Type of Adjuvant:**  NR  **Quantity of Adjuvant:**  N/A | **Intradermal**  **Sample Size (% Female):**  n=636 (57)  **Age (Mean ± SD):**  73.1 ± 6  **Dose:**  15µg  **Intradermal Administration Technique:**  NR | **Analysis Type:**  PP  **Maximum Follow-up:**  28 days  **Outcomes:**   - Adverse events - Serious adverse events - GMT - MFI - Seroconversion - Seroprotection |
|  |  |  |  | **Intradermal**  **Sample Size (% Female):**  n=627 (55)  **Age (Mean ± SD):**  72.9 ± 5.9  **Dose:**  21µg  **Intradermal Administration Technique:**  NR |  |
|  |  |  |  | **Intramuscular**  **Sample Size (% Female):**  n=317 (55)  **Age (Mean ± SD):**  73.4 ±5.9  **Dose:**  15µg |  |
|  |  |  |  | **Intramuscular**  **Sample Size (% Female):**  n=317 (57)  **Age (Mean ± SD):**  73 ± 6  **Dose:**  60µg |  |
| **Van Damme**  **et al.**  France, Belgium  (2010)^43^  Date of Recruitment:  Oct – Dec 2007  Trial Setting:  Multicenter | **Inclusion:**  65 years or older  **Exclusion:**  Fever, thrombocytopenia, chronic illness, congenital/acquired immunodeficiency, blood transfusion in past 3 months, alcohol or drug addiction, vaccination in past 4 weeks, influenza vaccination in last 6 months | **Total Sample Size (% Female):**  n= 795 (53.5)  **Age Range:**  ≥65 years  **Pre-existing Condition:**  None | **Vaccine Type:**  Trivalent  **Vaccine Characteristics:**  A/H1N1 (A/Solomon Islands/3/2006), A/H3N2 (  A/Wisconsin/67/2005), B Strain (B/Malaysia/2506/2004)  **Type of Adjuvant:**  MF59  **Quantity of Adjuvant:**  N/A | **Intradermal**  **Sample Size (% Female):**  n=395 (NR)  **Age (Mean ± SD):**  NR  **Dose:**  15µg  **Intradermal Administration Technique:**  NR | **Analysis Type:**  PP  **Maximum Follow-up:**  21 days  **Outcomes:**   - Adverse events - GMT - MFI - Seroconversion - Seroprotection |
|  |  |  |  | **Intramuscular**  **Sample Size (% Female):**  n=395 (NR)  **Age (Mean ± SD):**  NR  **Dose:**  15µg |  |
| **Non-Randomized** | | | | | |
| **Puig Barbera**  **et al.**  Spain  (2014)^46^  Date of Recruitment:  Oct – Nov 2011  Trial Setting:  Multicenter | **Inclusion:**  All community-dwelling adults aged ≥65 years as of 1 October2011, residing in Valencia Autonomous Community, Spain, and who were vaccinated against influenza during the 2011–2012influenza  **Exclusion:**  Admissions in the30 days following hospital discharge, duplicate cases (if the patient had more than one case admission, only the first was included), and institutionalized recipients of the split trivalent non-adjuvanted vaccine (Gripavac®, Sanofi-Pasteur MSD, Lyon, France). | **Total Sample Size (% Female):**  n= 164,021 (55.3)  **Age Range:**  ≥65 years  **Pre-existing Condition:**  NR | **Vaccine Type:**  Trivalent  **Vaccine Characteristics:**  A/H1N1 (A/California/7/2009), A/H3N2 (A/Perth/16/2009), B Strain (B/Brisbane/60/2008)  **Type of Adjuvant:**  NR  **Quantity of Adjuvant:**  N/A | **Intradermal**  **Sample Size (% Female):**  n=101,963 (NR)  **Mean Age (Range):**  72 (65-86)  **Dose:**  15µg  **Intradermal Administration Technique:**  NR | **Puig Barbera**  **et al.**  Spain  (2014)  Trial Design: Cohort  Date of Recruitment:  Oct – Nov 2011  Trial Setting:  Multicenter |

### Appendix C: Quality Assessment

Figure C1. Quality Assessment of Randomized Controlled Trials

| Author (Year) | Domain 1 | Domain 2 | Domain 3 | Domain 4 | Domain 5 | Overall Bias |
| --- | --- | --- | --- | --- | --- | --- |
| Ansaldi et al.  (2013)^18^ | Some Concern | Some Concern | Low  Concern | Low  Concern | Some Concern | Some Concern |
| Ansaldi et al.  (2012)^17^ | Some Concern | Some Concern | Low  Concern | Low  Concern | Some Concern | Some Concern |
| Arnou et al.  (2010)^19^ | Low  Concern | Some Concern | Low  Concern | Low  Concern | Some Concern | Some Concern |
| Boonak et al.  (2017)^20^ | Some Concern | Some Concern | Low  Concern | Low  Concern | Some Concern | Some Concern |
| Camiloni et al.  (2014)^21^ | Some Concern | Some Concern | Low  Concern | Low  Concern | Some Concern | Some Concern |
| Carter et al.  (2019)^22^ | Low  Concern | Low  Concern | Low  Concern | Low  Concern | Some Concern | Some Concern |
| Chan et al.  (2014)^23^ | Low  Concern | Some Concern | Low  Concern | Low  Concern | Some Concern | Some Concern |
| Chi et al.  (2010)^24^ | Some Concern | Some Concern | Low  Concern | Low  Concern | Some Concern | Some Concern |
| Chuaychoo et al.  (2019)^25^ | Some Concern | Some Concern | Low  Concern | Low  Concern | Some Concern | Some Concern |
| Chuaychoo et al.  (2016)^26^ | Some Concern | Some Concern | Low  Concern | Low  Concern | Some Concern | Some Concern |
| Chuaychoo et al.  (2010)^27^ | High  Concern | Some Concern | Low  Concern | Low  Concern | Some Concern | High  Concern |
| Della Cioppa et al.  (2014)^28^ | Some Concern | Some Concern | Low  Concern | Low  Concern | Some Concern | Some Concern |
| Esposito et al.  (2011)^29^ | Low  Concern | Some Concern | Low  Concern | Low  Concern | Some Concern | Some Concern |
| Frenck et al.  (2011)^30^ | Some Concern | Some Concern | Low  Concern | Low  Concern | Some Concern | Some Concern |
| Garg et al.  (2016)^31^ | Low  Concern | Some Concern | Low  Concern | Low  Concern | Some Concern | Some Concern |
| Gorse et al.  (2013)^32^ | Some Concern | Some Concern | Low  Concern | Low  Concern | Some Concern | Some Concern |
| Han et al.  (2013)^33^ | High  Concern | Some Concern | Low  Concern | High  Concern | Some Concern | High  Concern |
| Hung et al.  (2016)^34^ | Low  Concern | Low  Concern | Low  Concern | Low  Concern | Some Concern | Some Concern |
| Hung et al.  (2014)^36^ | Some Concern | Some Concern | Low  Concern | Low  Concern | Some Concern | Some Concern |
| Hung et al.  (2012)^35^ | Some Concern | Some Concern | Low  Concern | Low  Concern | Some Concern | Some Concern |
| Leung et al.  (2017)^37^ | Some Concern | Some Concern | Low  Concern | Low  Concern | Some Concern | Some Concern |
| Levin et al.  (2016)^45^ | Some Concern | Some Concern | Low  Concern | Low  Concern | Some Concern | Some Concern |
| Levin et al.  (2014)^44^ | Some Concern | Some Concern | Low  Concern | Low  Concern | Some Concern | Some Concern |
| Nougaredet al.  (2014)^38^ | Some Concern | Some Concern | Low  Concern | Low  Concern | Some Concern | Some Concern |
| Patel et al.  (2010)^39^ | Some Concern | Some Concern | Low  Concern | Low  Concern | Some Concern | Some Concern |
| Seo et al.  (2014)^40^ | Some Concern | Some Concern | Low  Concern | Low  Concern | Some Concern | Some Concern |
| Song et al.  (2013)^41^ | Some Concern | Some Concern | Low  Concern | Low  Concern | Some Concern | Some Concern |
| Tsang et al.  (2014)^42^ | Some Concern | Some Concern | Low  Concern | Low  Concern | Some Concern | Some Concern |
| Van Damme et al.  (2010)^43^ | Some Concern | Some Concern | Low  Concern | Low  Concern | Some Concern | Some Concern |

### Appendix D: Forest Plots

Figure D1. Seroconversion 3 mcg ID vs 15 mcg IM

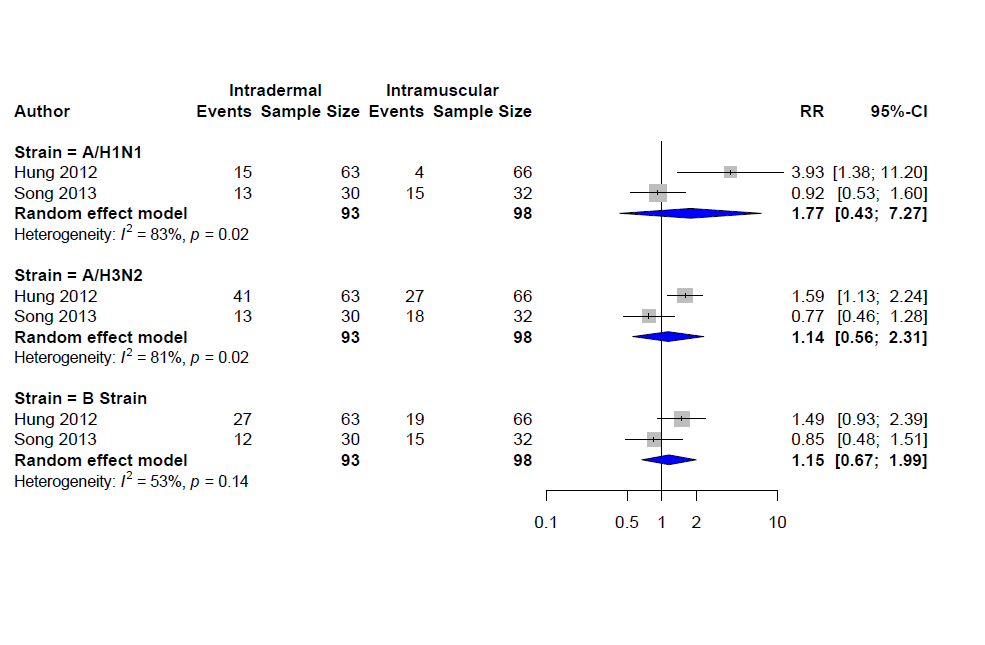

Figure D2. Seroconversion 6 mcg ID vs 15 mcg IM (* Inflexal vs Fluad Brand)

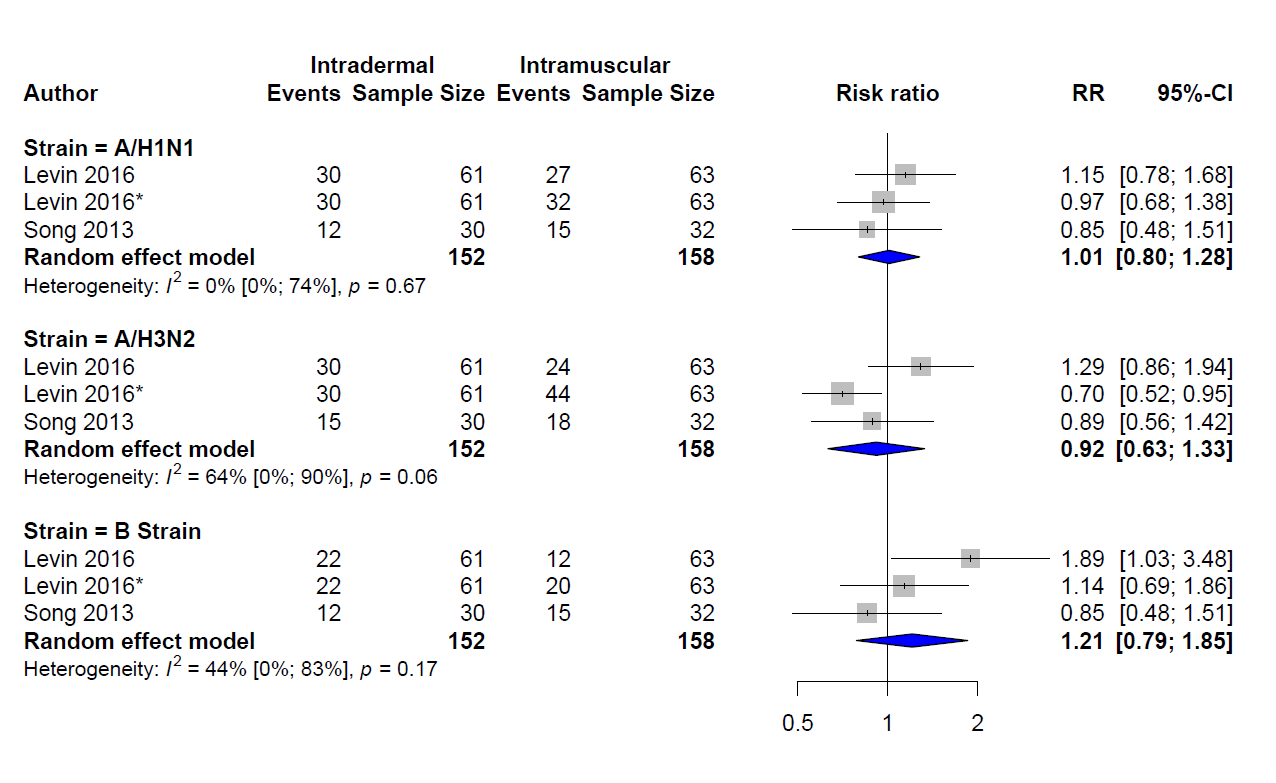

Figure D3. Seroconversion 7.5 mcg ID vs 15 mcg IM (* Inflexal vs Fluad)

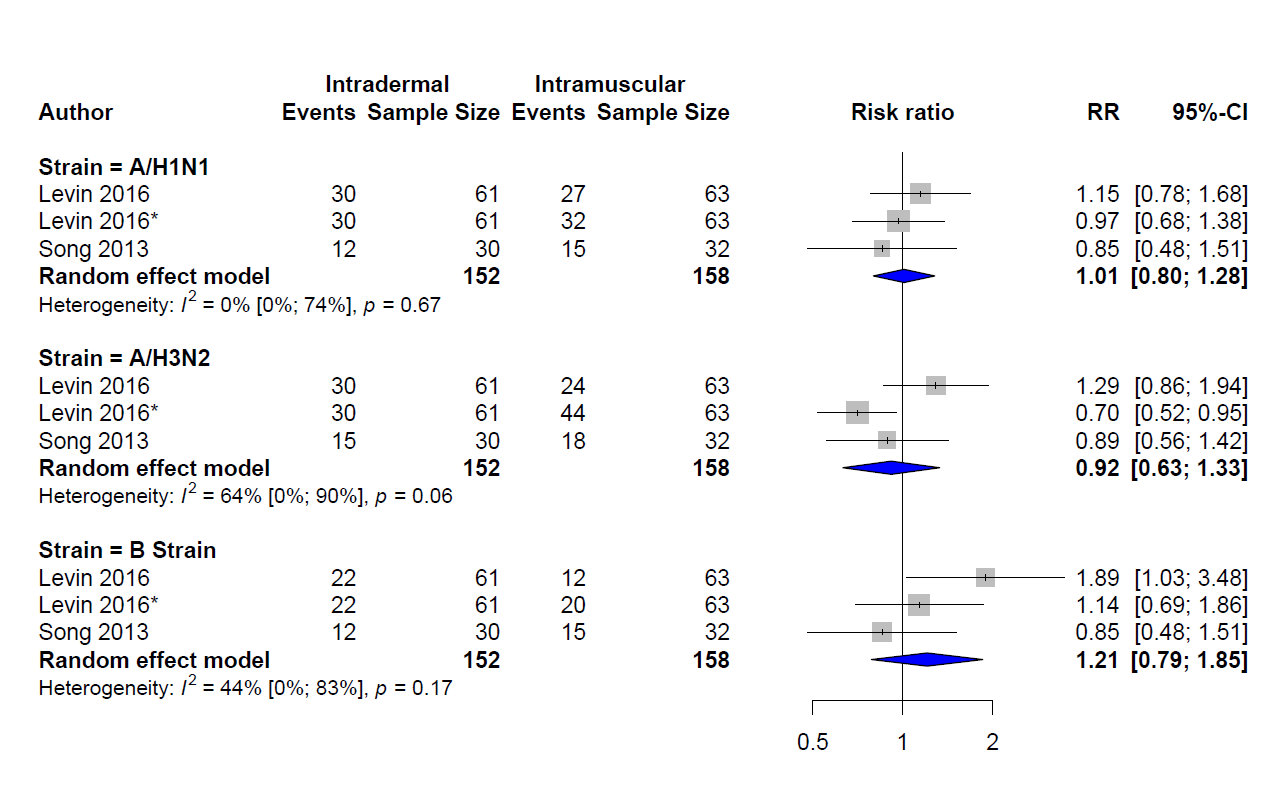

Figure D4. Seroconversion 9 mcg ID vs 15 mcg IM in All Population: † Soluvia Microneedle

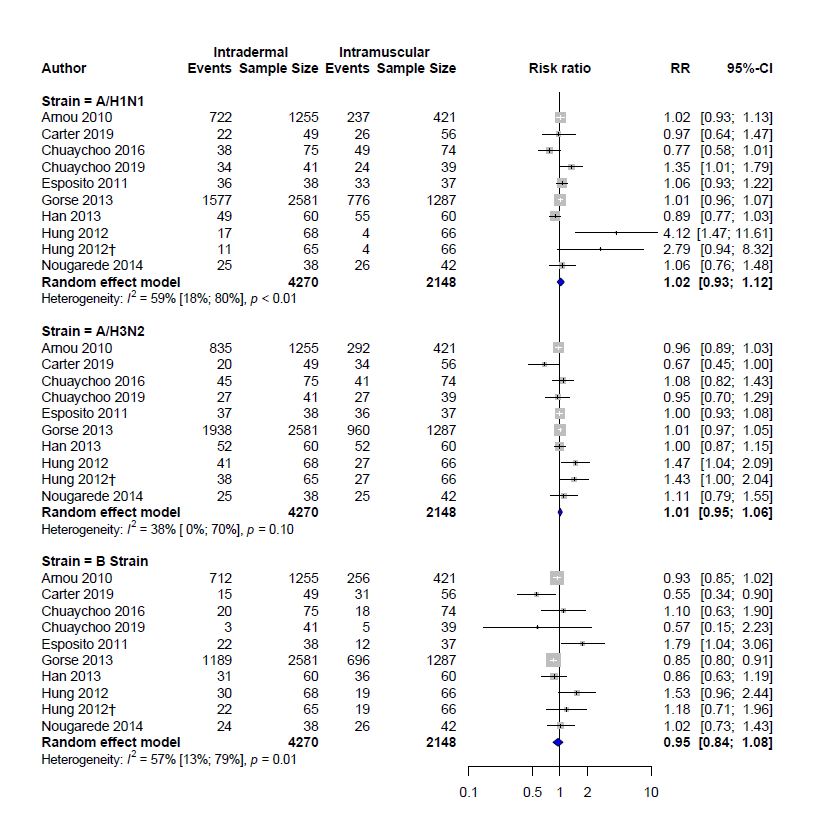

Figure D5. Seroconversion in Elderly 9 mcg ID vs 15 mcg IM

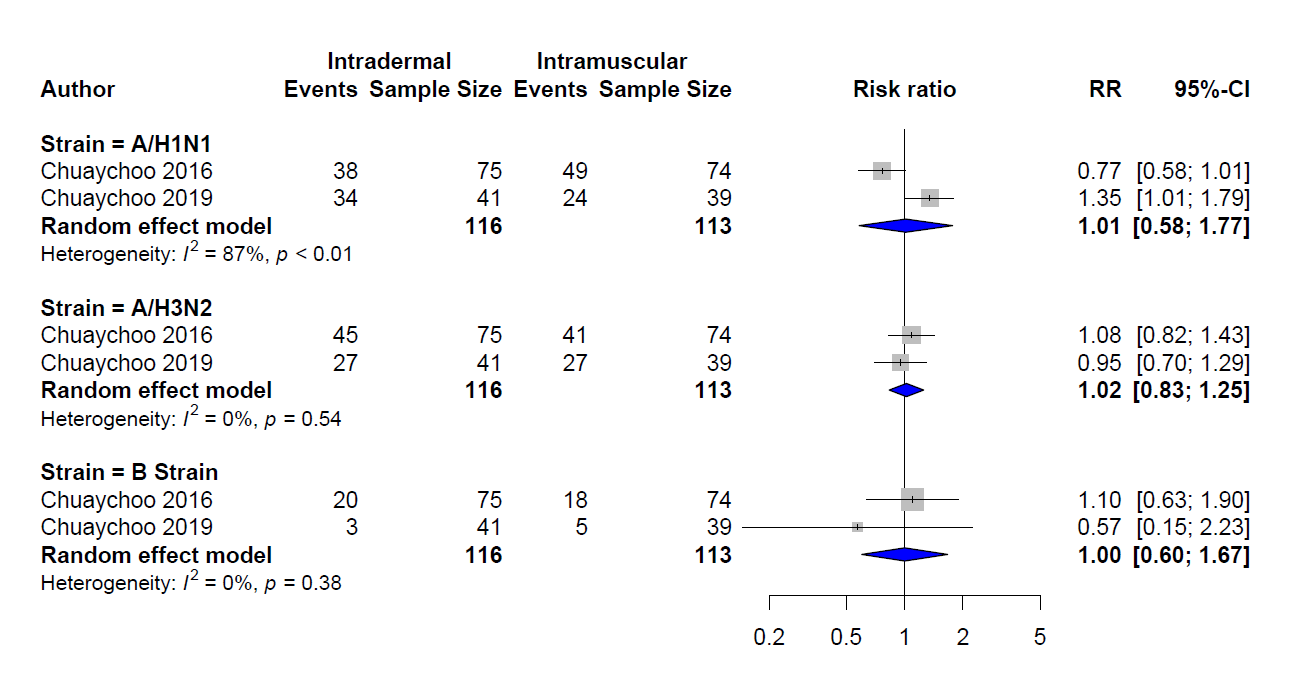

Figure D6. Seroconversion 15 mcg ID vs 15 mcg IM (*Inflexa vs Inflexa, ** Intanza vs Inflexa,†Inflexa vs Fluad, ‡Intanza vs Fluad, ᵻ Adjuvanted)
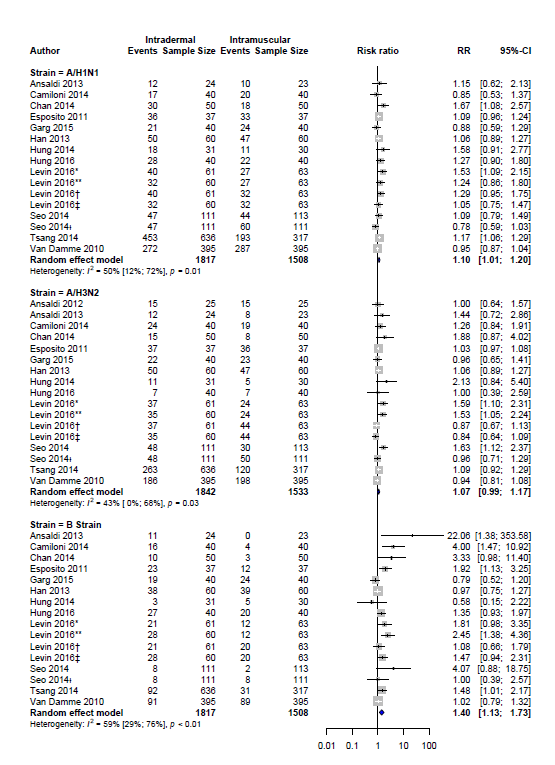

Figure D7. Seroconversion in the Elderly15 mcg ID vs 15 mcg IM (*Inflexa vs Inflexa, ** Intanza vs Inflexa,†Inflexa vs Fluad, ‡Intanza vs Fluad, ᵻ Adjuvanted)

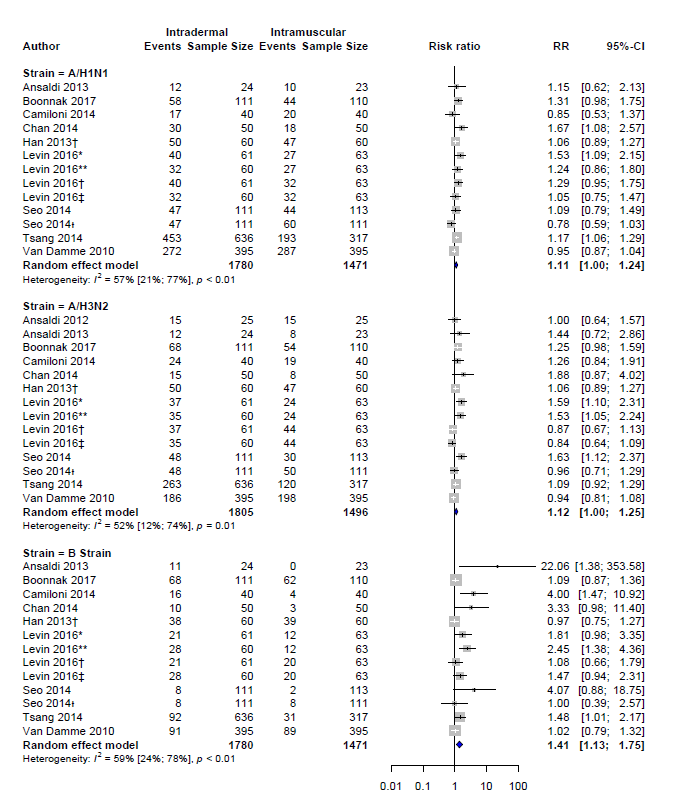

Figure D8. Seroprotection 3 mcg ID vs 15 mcg IM

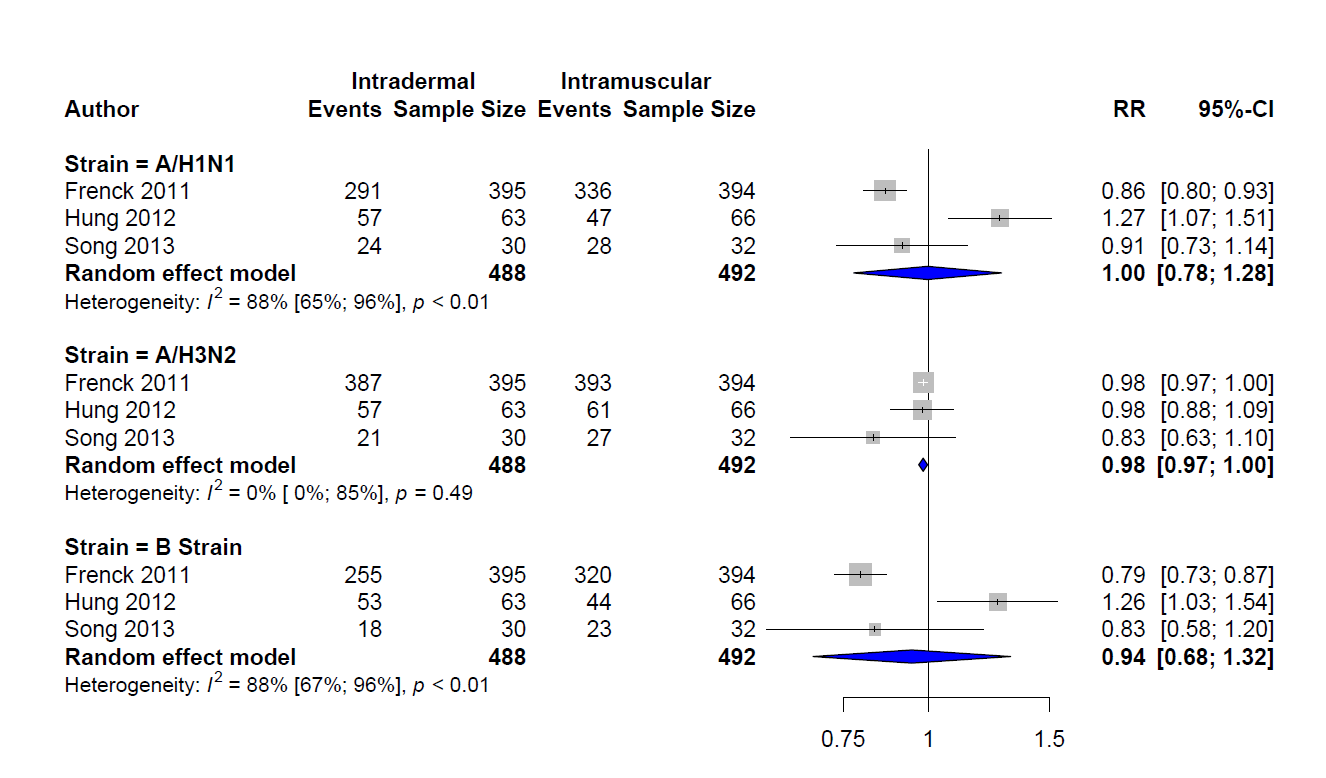

Figure D9. Seroprotection 6 mcg ID vs 15 mcg IM

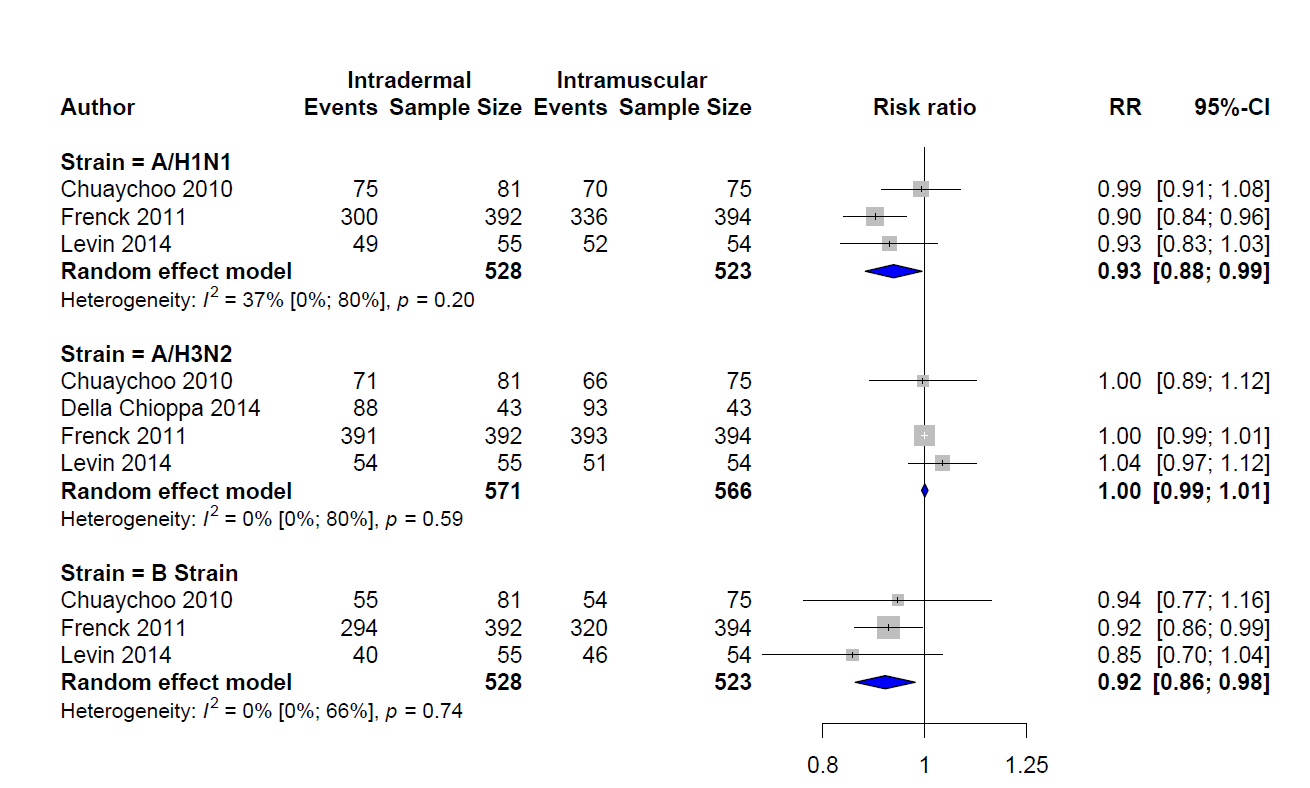

Figure D10. Seroprotection 7.5 mcg ID vs 15 mcg IM (*Inflexa vs Fluad Brand)

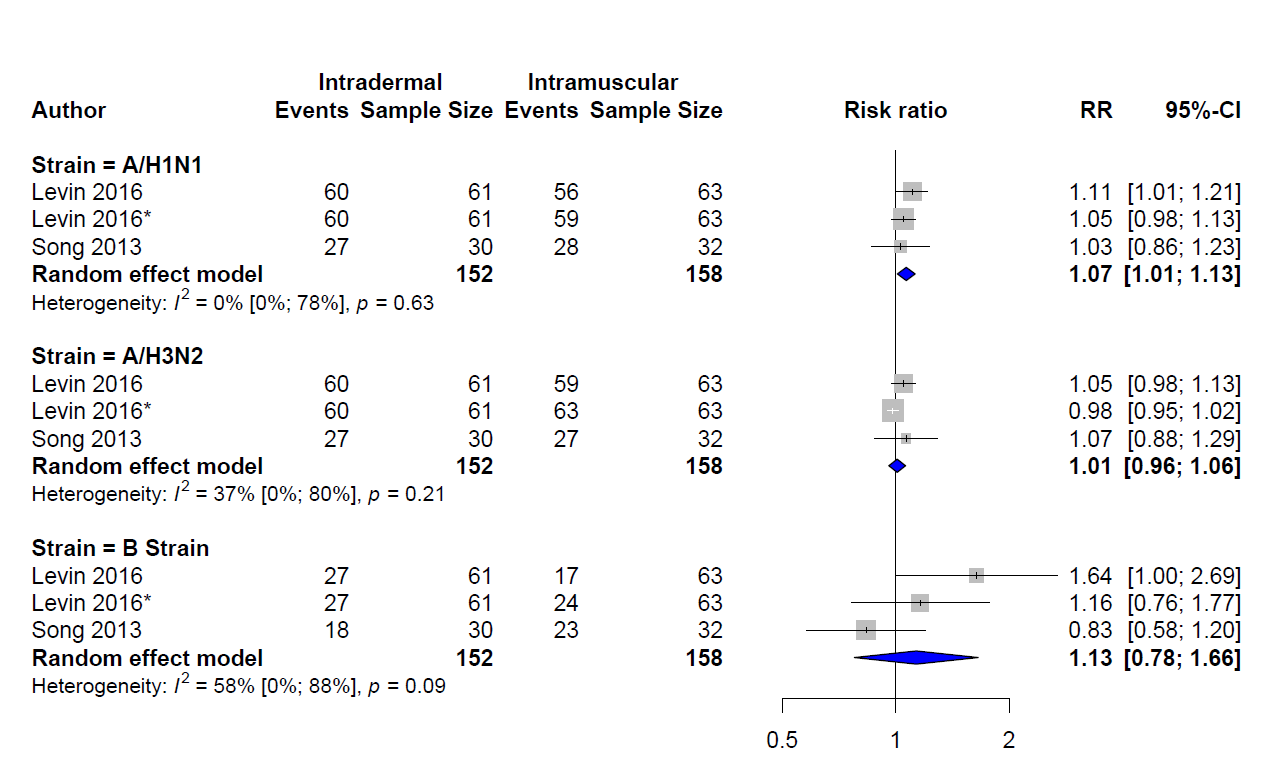

Figure D11. Seroprotection 9 mcg ID vs 15 mcg IM († Soluvia Microneedle, * Two Injections)

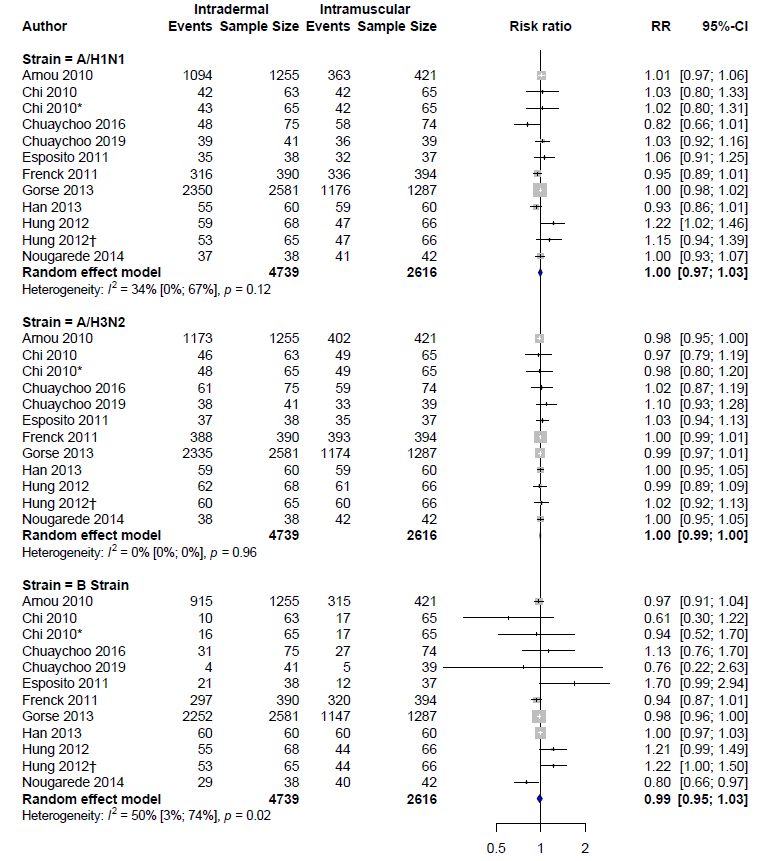

Figure D12. Seroprotection Elderly 9 mcg ID vs 15 mcg IM: ᵻᵻ Two Injections

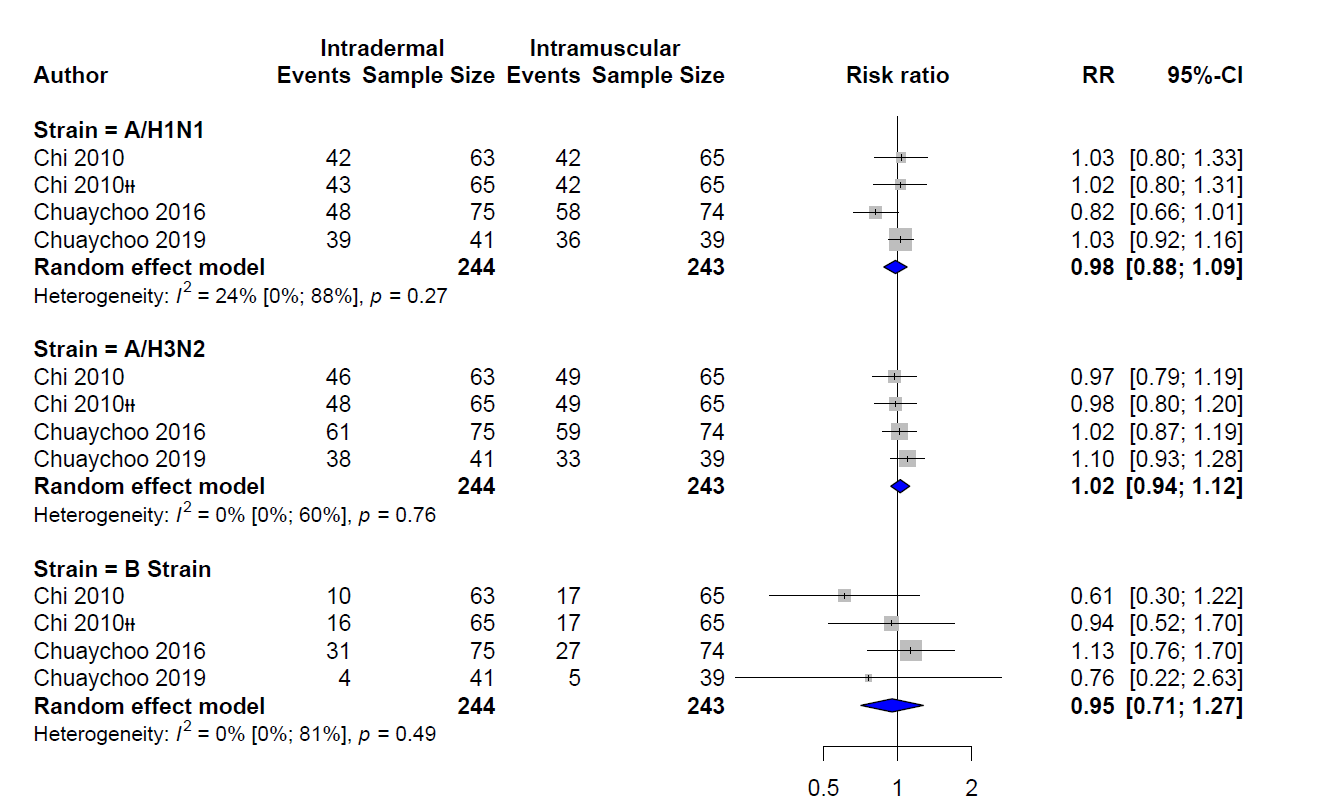

Figure D13. Seroprotection 15 mcg ID vs 15 mcg IM (*Inflexa vs Inflexa, ** Intanza vs Inflexa,†Inflexa vs Fluad, ‡Intanza vs Fluad, ᵻ Adjuvanted)
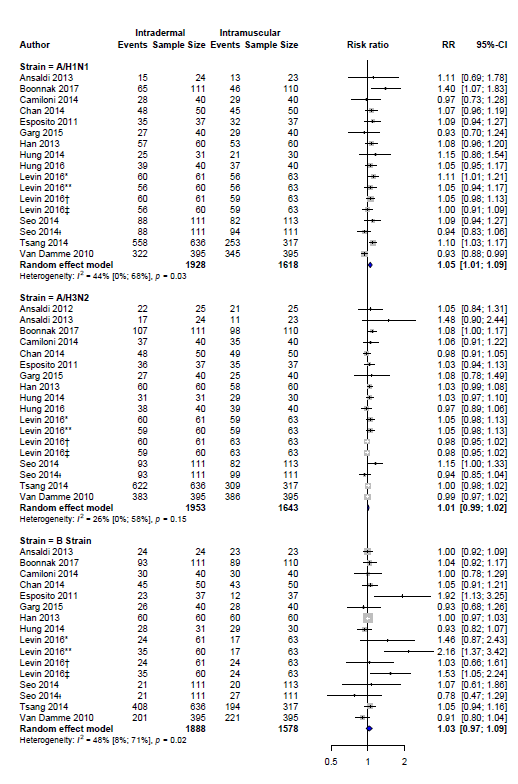

Figure D14. Seroprotection in Elderly 15 mcg ID vs 15 mcg IM (*Inflexa vs Inflexa, ** Intanza vs Inflexa,†Inflexa vs Fluad, ‡Intanza vs Fluad, ᵻ Adjuvanted)

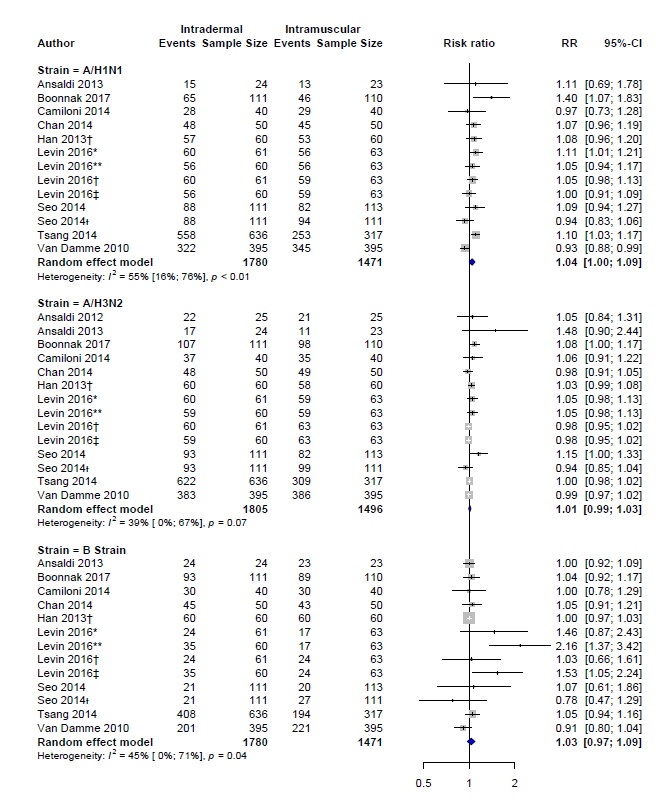

Figure D15. GMT 3 mcg ID vs 15 mcg IM

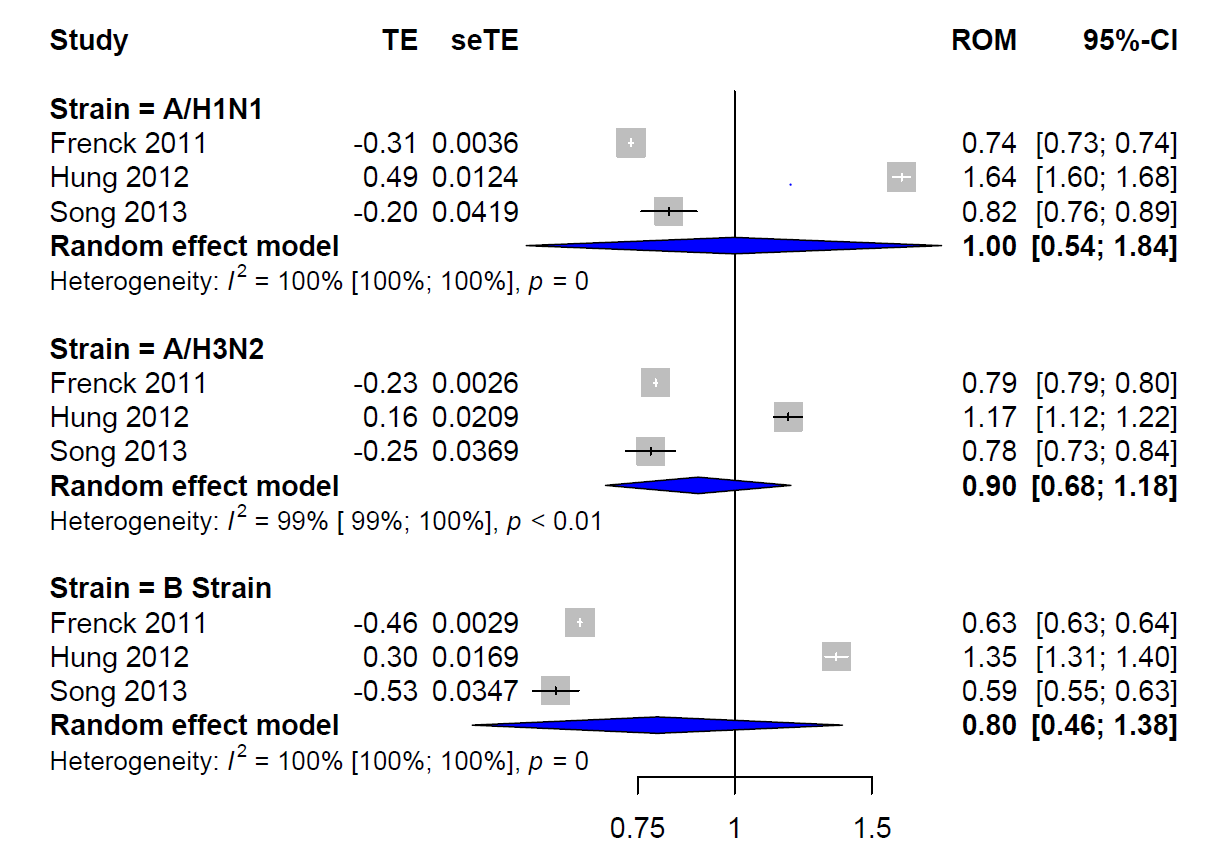

Figure D16. GMT 6 mcg ID vs 15 mcg IM

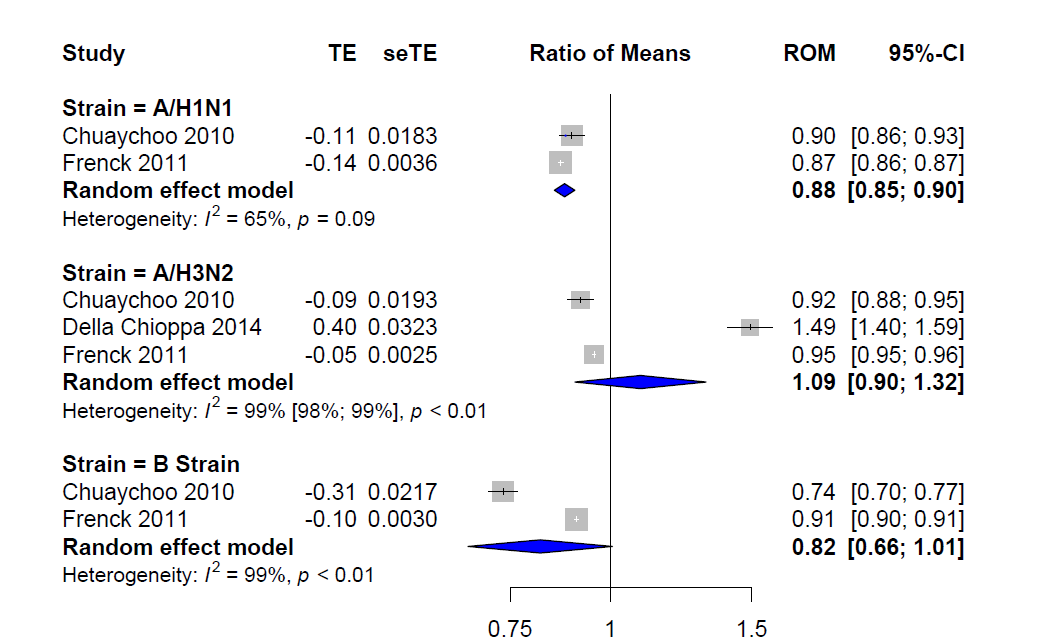

Figure D17. GMT 9 mcg ID vs 15 mcg IM (* Two Doses, †Soluvia Device)

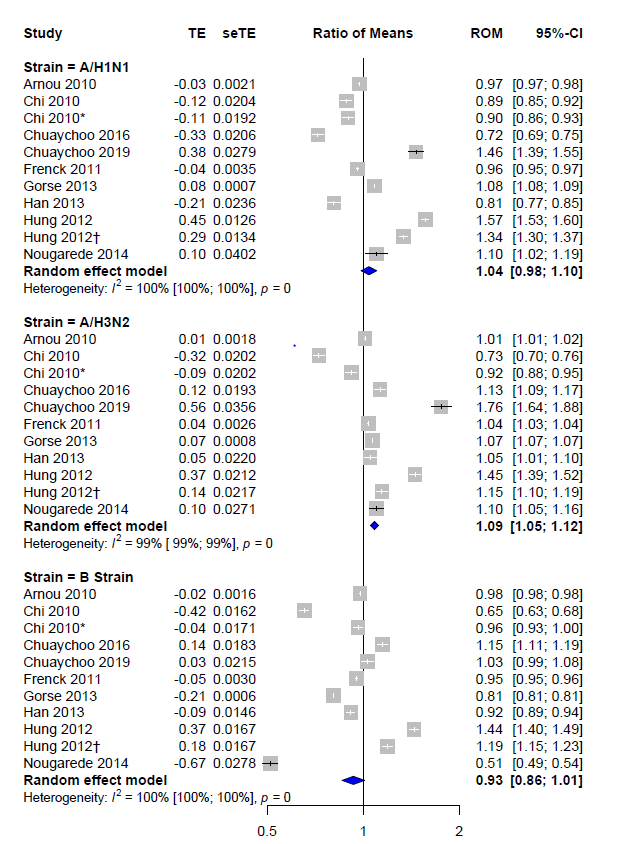

Figure D18. GMT in Elderly 9 mcg ID vs 15 mcg IM (ᵻᵻ Two Doses)

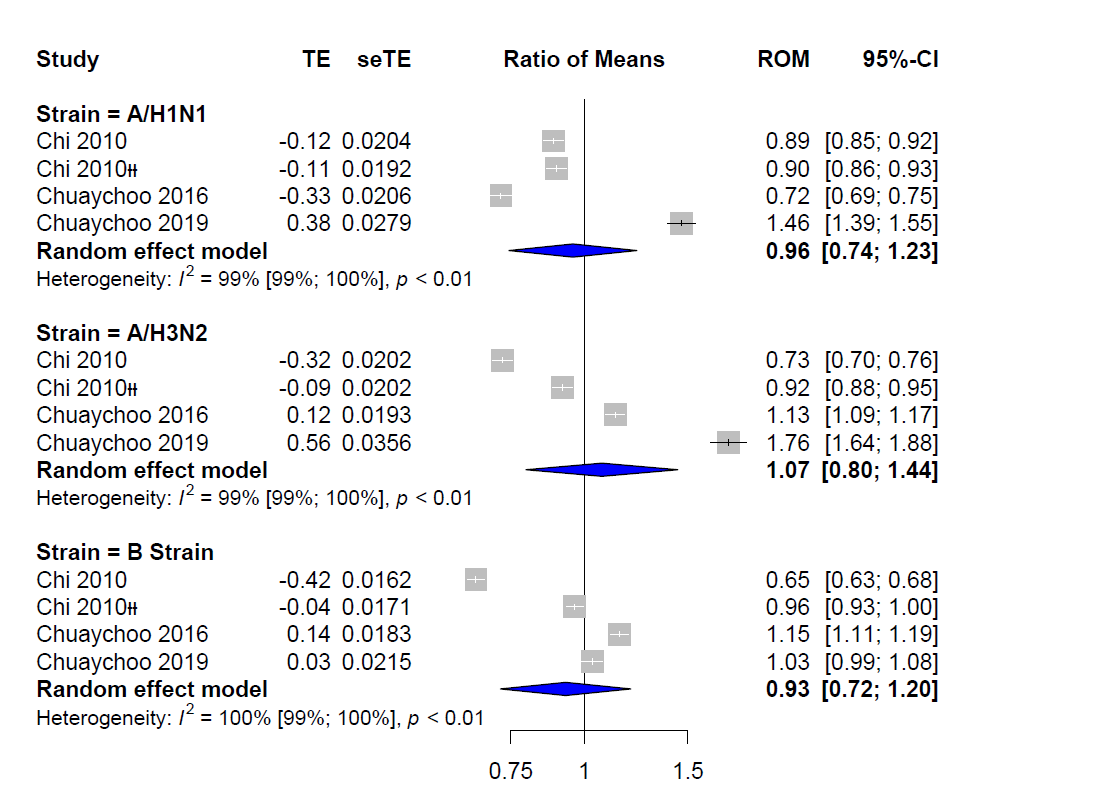

Figure D19. GMT 15 mcg ID vs 15 mcg IM (ᵻ Adjuvanted)

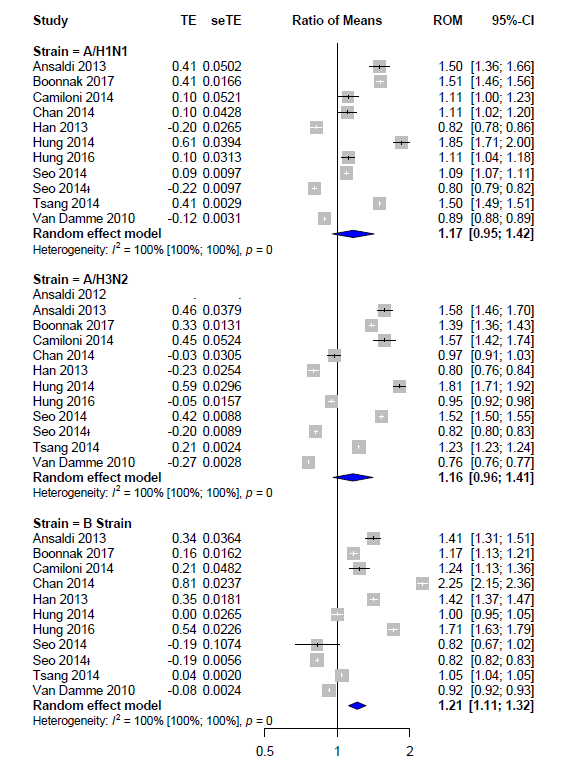

Figure D20. GMT in Elderly 15 mcg ID vs 15 mcg IM(ᵻ Adjuvanted, †≥60 Years Old)

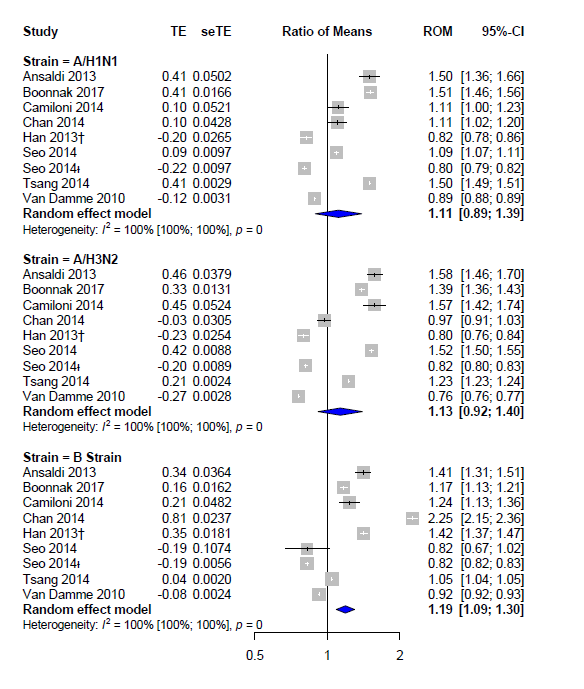

Figure D21. Local Adverse Events 3 mcg ID vs 15 mcg IM

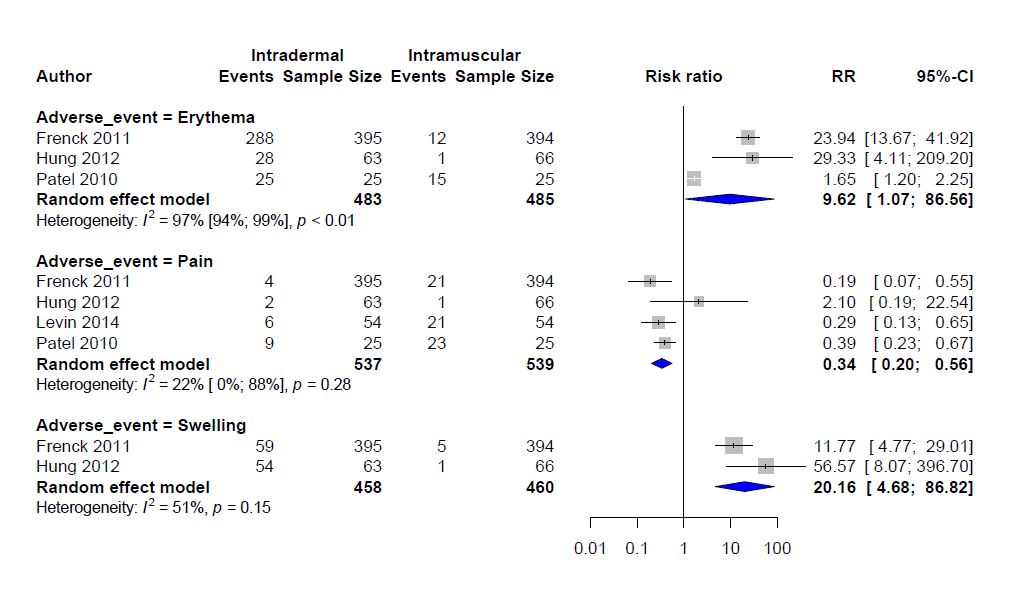

Figure D22. Local Adverse Events 6 mcg ID vs 15 mcg IM

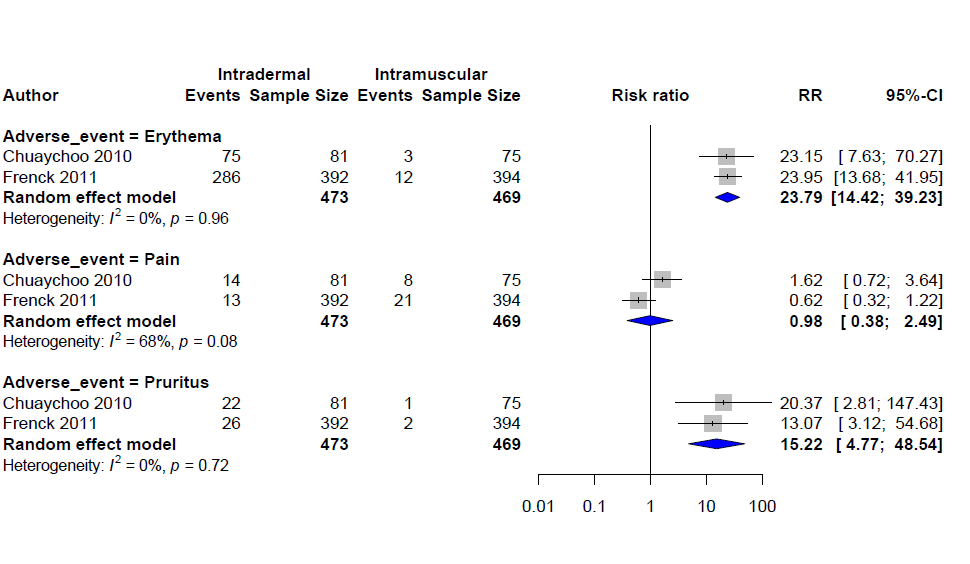

Figure D23. Local Adverse Events 9 mcg ID vs 15 mcg IM (††Two Doses, ‡Soluvia Device)

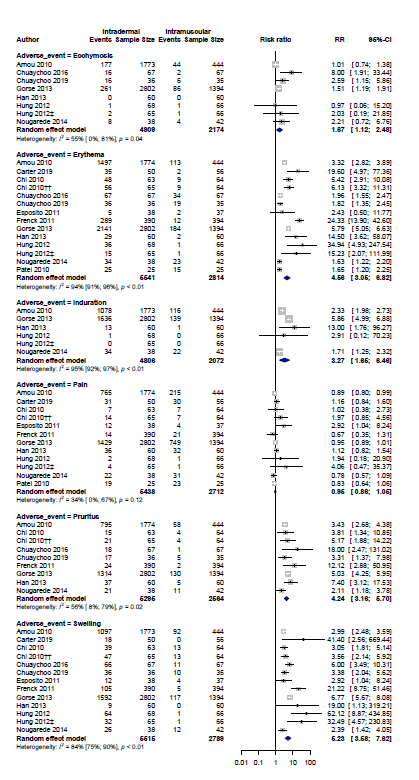

Figure D24. Local Adverse Events 15 mcg ID vs 15 mcg IM(*Inflexa vs Inflexa, ** Intanza vs Inflexa,†Inflexa vs Fluad, ‡Intanza vs Fluad, ᵻ Adjuvanted)

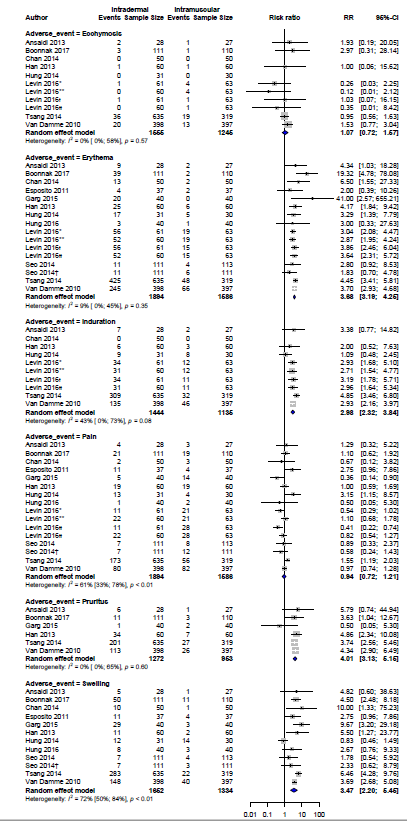

Figure D25. Systemic Adverse Events 3 mcg ID vs 15 mcg IM

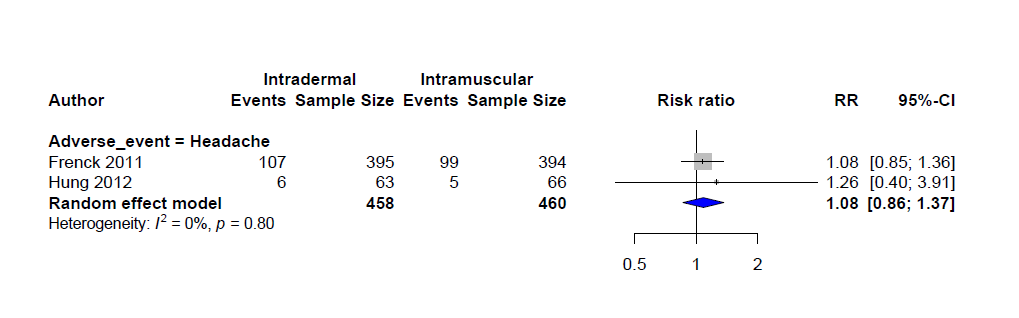

Figure D26. Systemic Adverse Events 6 mcg ID vs 15 mcg IM

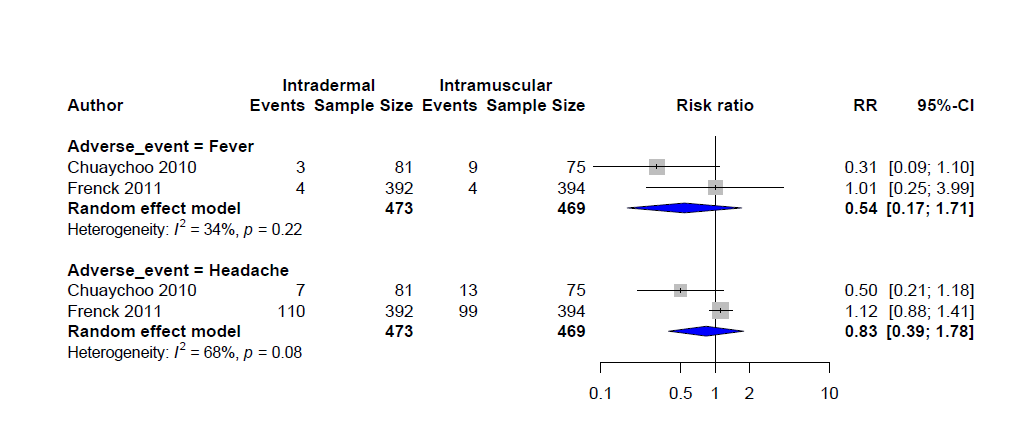

Figure D27. Systemic Adverse Events 9 mcg ID vs 15 mcg IM (‡ Soluvia, ††Two Doses)

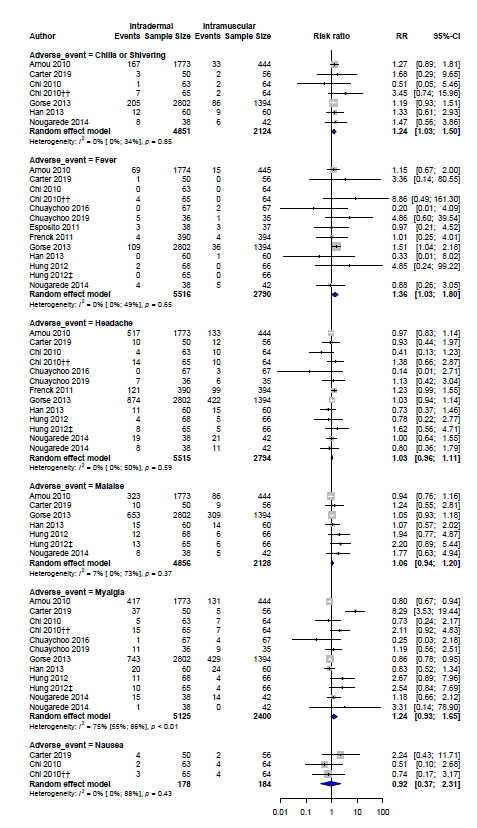

Figure D28. Systemic Adverse Events 15 mcg ID vs 15 mcg IM (*Inflexa vs Inflexa, ** Intanza vs Inflexa,†Inflexa vs Fluad, ‡Intanza vs Fluad, ᵻ Adjuvanted)

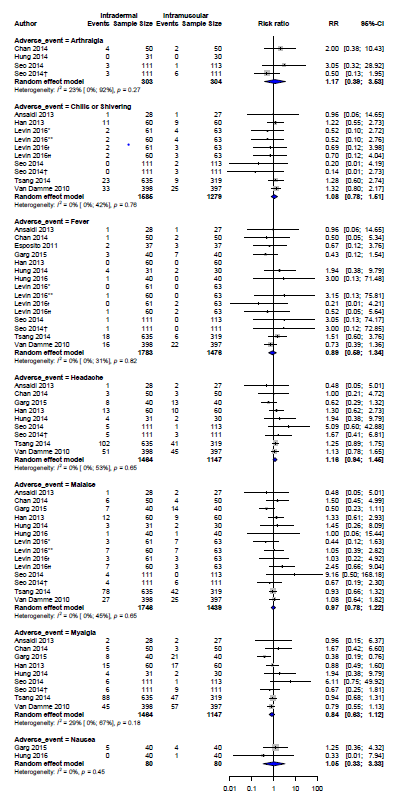

### Appendix E: Funnel Plots

Figure E1. Funnel Plot Seroconversion 9 mcg ID vs 15 mcg IM

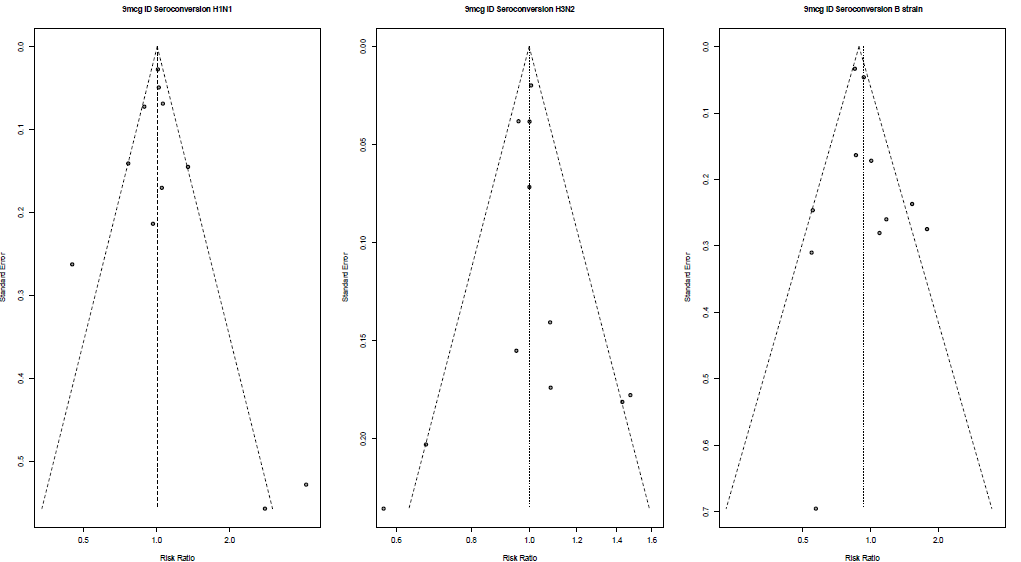

Figure E2. Funnel Plot Seroprotection 9 mcg

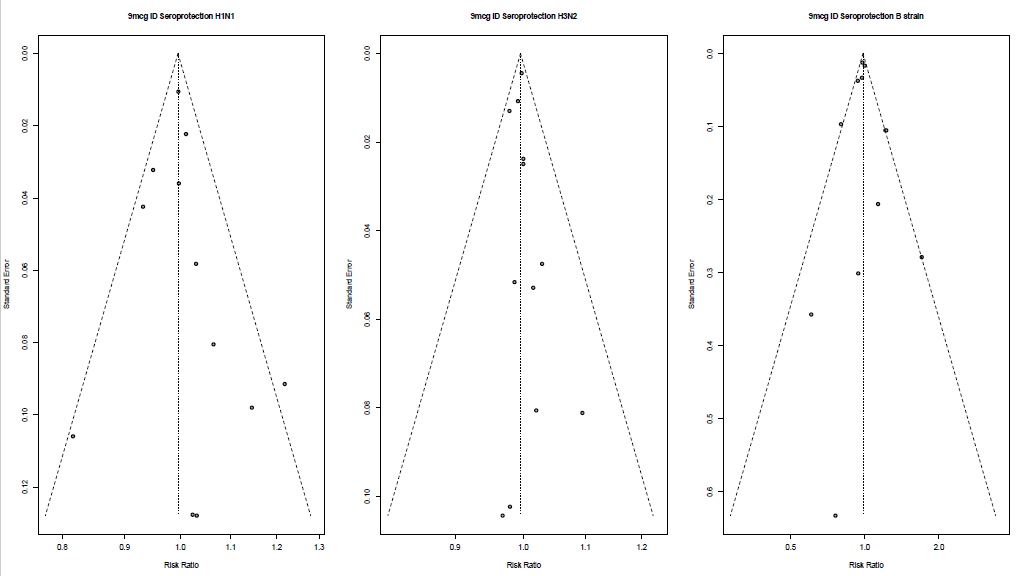

Figure E3. Funnel Plot GMT 9 mcg ID vs 15 mcg IM

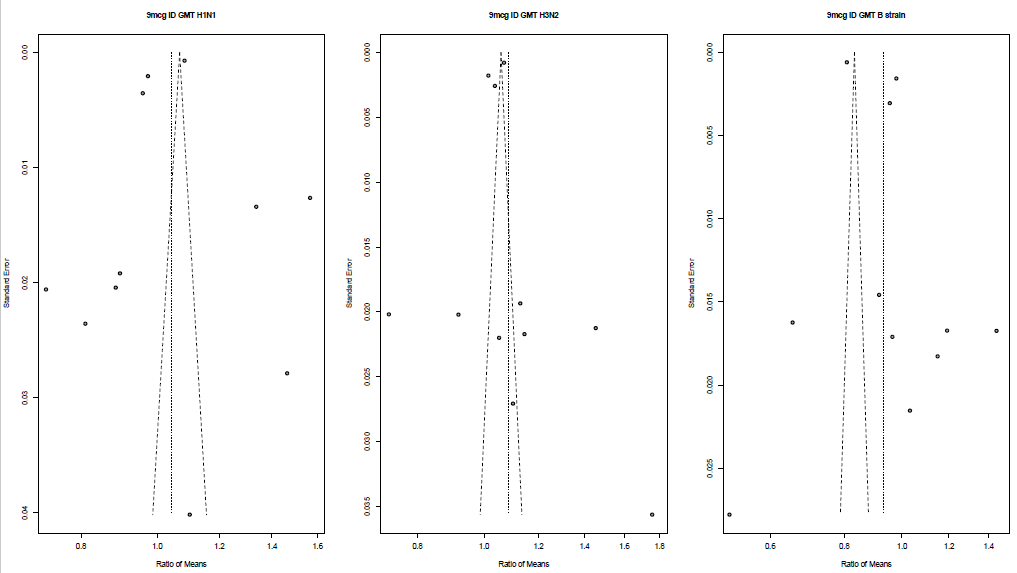

Figure E4. Funnel Plot Seroconversion 15 mcg ID vs 15 mcg IM

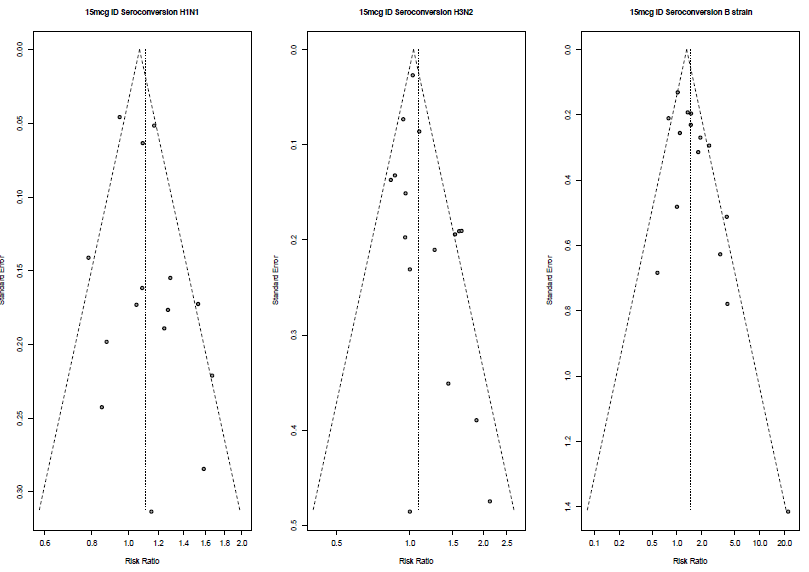

Figure E5. Funnel Plot Seroprotection 15 mcg ID vs 15 mcg IM

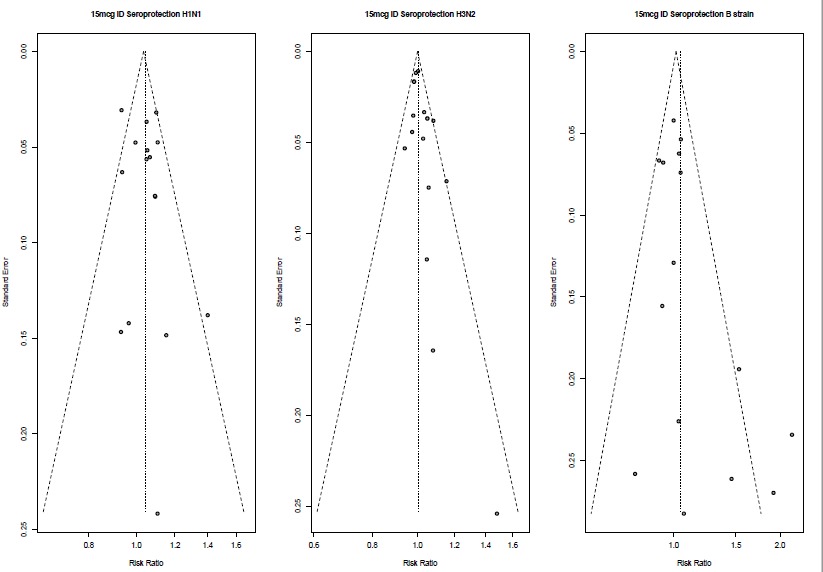

Figure E6. Funnel Plot GMT 15 mcg ID vs 15 mcg IM

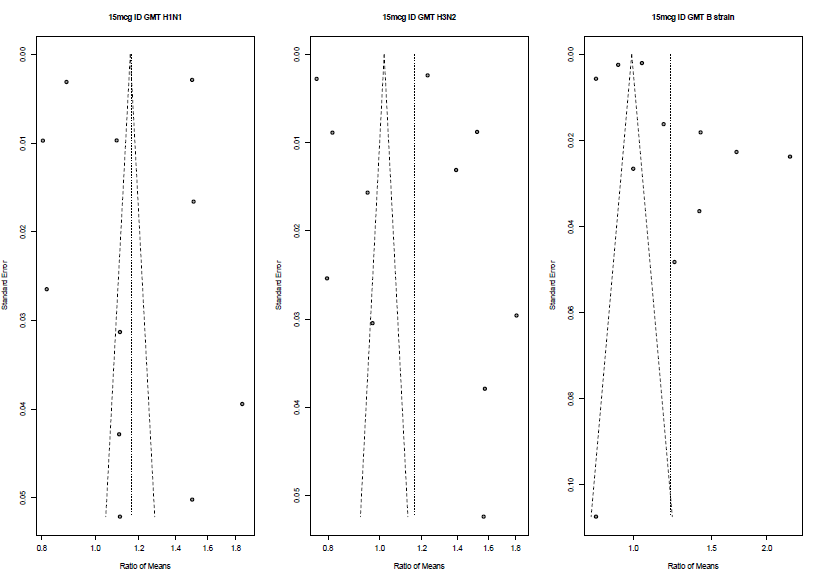
